# Systematic examination of T cell responses to SARS-CoV-2 versus influenza virus reveals distinct inflammatory profile

**DOI:** 10.1101/2020.08.27.20183319

**Authors:** Jaclyn C. Law, Wan Hon Koh, Patrick Budylowski, Jonah Lin, FengYun Yue, Kento T. Abe, Bhavisha Rathod, Melanie Girard, Zhijie Li, James M. Rini, Samira Mubareka, Allison McGeer, Adrienne K. Chan, Anne-Claude Gingras, Tania H. Watts, Mario Ostrowski

## Abstract

There is a pressing need for an in-depth understanding of immunity to SARS-CoV-2. Here we investigated T cell recall responses to fully glycosylated Spike trimer, recombinant N protein as well as to S, N, M and E peptide pools in the early convalescent phase. All subjects showed SARS-CoV-2-specific T cell responses to at least one antigen. SARS-CoV-2-specific CD4+ T cells were primarily of the central memory phenotype and exhibited a lower IFN-γ to TNF-α ratio compared to influenza-specific responses of the same donors, independent of disease severity. SARS-CoV-2-specific T cells were less multifunctional than influenza-specific T cells, particularly in severe cases, potentially suggesting exhaustion. High IL-10 production was noted in response to N protein, possibly contributing to immunosuppression, with potential implications for vaccine design. We observed granzyme B+/IFN-γg+ CD4+ and CD8+ proliferative responses to peptide pools in most individuals, with CD4+ responses predominating over CD8+ responses. Peripheral T follicular helper responses to S or N strongly correlated with serum neutralization assays as well as RBD-specific IgA. Overall, T cell responses to SARS-CoV-2 are robust, however, CD4+ Th1 responses predominate over CD8+ responses and are more inflammatory with a weaker Tfh response than influenza-specific CD4+ responses, potentially contributing to COVID-19 disease.

---

The disease COVID-19, caused by the novel coronavirus (CoV), SARS-CoV-2, emerged in China in late 2019 and is currently causing a devastating pandemic (1-3). Despite the severity of the disease in some individuals, the vast majority of infected people recover, indicating that they have made an effective immune response that clears the virus. Moreover, studies in rhesus macaques demonstrate that SARS-CoV-2 induces protective immunity against rechallenge at least out to 35 days (4, 5). Adaptive immunity, mounted by T and B lymphocytes, is critical for clearance of viral infections and for protection against reinfection. Most studies to date show that people infected with SARS-CoV-2 produce Spike (S) and receptor binding domain (RBD) specific-IgG and neutralizing antibodies within two to four weeks of infection (6-13). Although some studies have suggested that antibody responses of people with mild or no symptoms can fall off rapidly (7, 14, 15), other studies suggest IgG responses are relatively stable over the first 3-4 months, with peak responses followed by a gradual decline as observed in a normal IgG response (13, 16, 17). In contrast, IgA responses to SARS-CoV-2 start early and decay rapidly (17). In the absence of complete virus neutralization, T cells are critical for eliminating virus-infected cells. Moreover, CD4+ T cell responses, and in particular T follicular helper (Tfh) responses, are critical for generation of high affinity long-lived antibody responses (18). Follow-up studies of the SARS-CoV-1 outbreak in 2003 showed that antibody responses fell off substantially between 3 and 5 years in most individuals (19), whereas T cell responses could be detected for more than 11 years (20). Moreover, nucleocapsid (N)-reactive T cells in SARS-CoV-1 recovered patients at 17 years post-infection showed substantial cross-reactivity to SARS-CoV-2 N peptides (21). Thus, T cells likely represent an important part of protective immunity to SARS-CoV-2.

Several studies have examined T cell responses to SARS-CoV-2 with most studies using restimulation with overlapping peptide pools from several SARS-CoV-2 open reading frames (21-28). Responses to restimulation with intact N, S-RBD domain and protease proteins have also been reported (11). The studies to date have used a variety of readouts to determine T cell specificity including activation markers, intracellular cytokine production, IFN-γ EliSpot or measurements of cytokines in the supernatants by multiplex assays. In general, the majority of confirmed SARS-CoV-2 cases have shown CD4+ and CD8+ T cell responses to SARS-CoV-2 antigens in the acute and early convalescent phase, dominated by a Th1 response with some studies also reporting Th2 or Th17 responses (reviewed in (29)). CD8+ T cell responses have also been detected in the majority of, but not all, donors. There is also evidence of cross-reactive T cells in 20-50% of donors who donated blood pre-pandemic. These cross-reactive responses are to peptides conserved between seasonal coronaviruses and SARS-CoV-2 (21, 22, 28, 30, 31).

Given the consistent findings of Th1 and CD8+ T cell responses in the acute and early convalescent stage of SARS-CoV2, often with the strongest responses detected in the more severe cases, it is not yet clear why the immune system fails to rapidly control the virus in some patients. Here we undertook a systematic functional examination of T cell responses to SARS-CoV-2 in a cohort of 13 SARS-COV-2 recovered individuals with a range of disease severity who provided leukapheresis samples in the early convalescent phase (4-12 weeks). Specifically, we examined T cell phenotype, cytokine production, and proliferation to SARS-CoV-2 proteins and peptides and compared them to seasonal influenza responses. We also identified peripheral T follicular (pTfh) IL-2 producing CCR7+CXCR5+ cells in response to SARS-CoV-2 antigens in some donors and found that the frequency of these cells strongly correlated with serum neutralization assays and RBD-specific IgA but were less frequent than those observed in response to influenza. Our study reveals new insights into the recall response of SARS-Cov-2 in the early convalescent phase, highlighting that SARS-CoV-2-specific CD4+ T cell responses are more inflammatory and show a weaker pTfh response than influenza A-specific CD4+ recall responses within the same donors.

## Results

### Patient characteristics

Thirteen COVID-19 convalescent donors who had recently tested positive for SARS-CoV-2 by PCR, and a single SARS-CoV-1 patient from 2003 were consented for leukapheresis to obtain plasma and PBMC (**Table 1)**. Samples for SARS-CoV-2 convalescent individuals were collected from 27 days to 90 days post onset of symptoms. Disease severity ranged from asymptomatic, mild (non-hospitalized), moderate (hospitalized not ICU) to severe (ICU). The average age was 53 (range 31-72), and 8 out of 13 were male (**Table I**).

**Table I.**
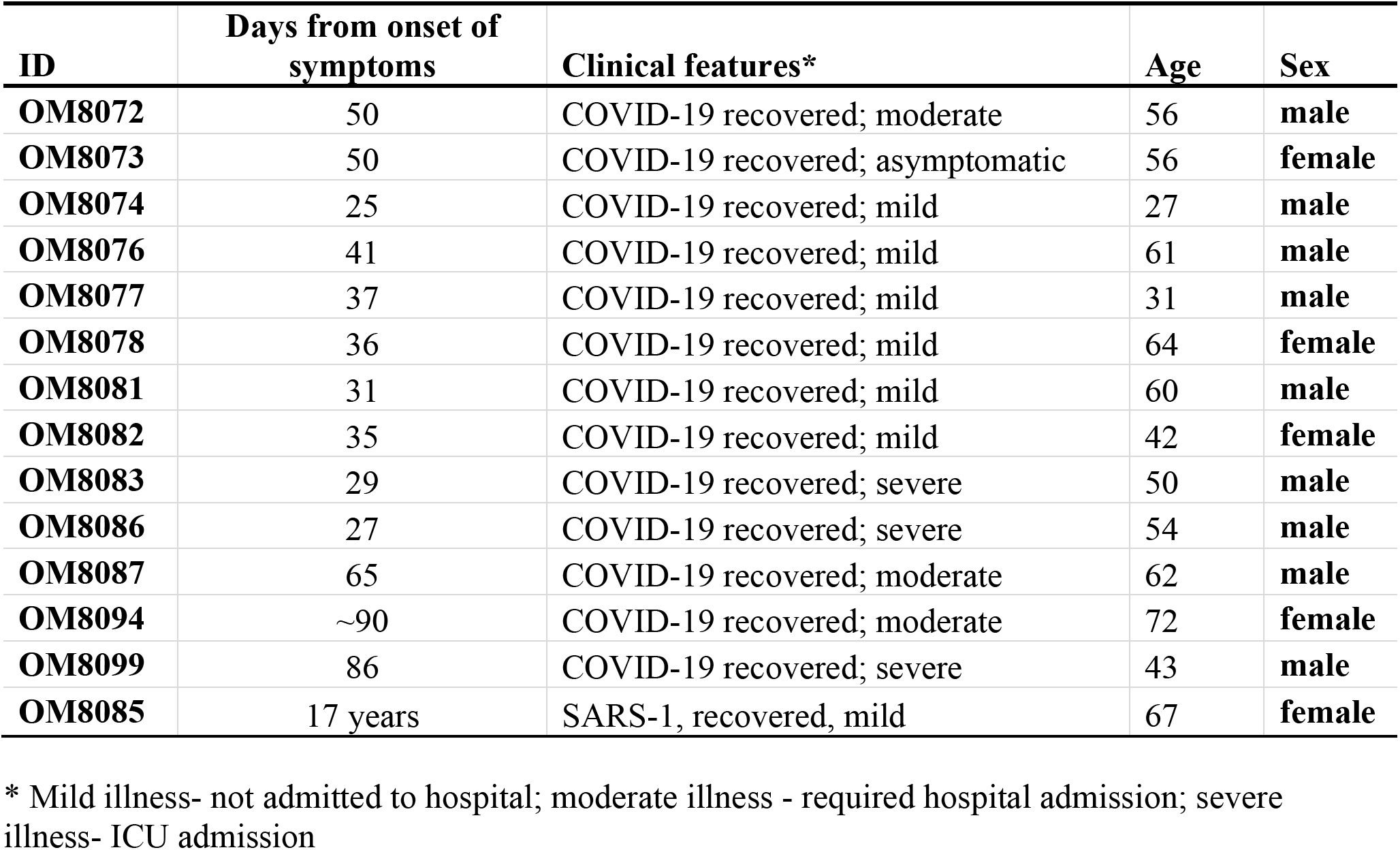
Clinical Characteristics of Participants.

### Ex vivo intracellular cytokine responses to Spike and N proteins in convalescent COVID-19 patients

To date, most studies have used overlapping peptide pools to assess antigen-specific T cell responses to SARS-CoV-2. Here we used intact glycosylated S from SARS-CoV-2 and seasonal human CoV-OC43 (OC43), as well as recombinant *E. coli* expressed SARS-CoV-2 N to determine how T cells respond functionally to SARS-CoV-2 under conditions where antigen processing is required. Intracellular cytokine staining (ICC) of ex vivo PBMC was conducted to determine the frequency of IFN-γ, TNF-α and IL-2-producing cells, with the gating strategy shown in **Figure S1A**. Following 18hrs of stimulation, with GolgiStop and GolgiPlug added for the last 6 hrs, SARS-CoV-2 S-specific CD4+ T cells were detected in 54% of donors based on IFN-γ production, 75% of donors based on TNF-α production and 85% of donors based on IL-2 production. N-specific CD4+ T cells were detected in 38% of donors based on IFN-γ, 58% based on TNF-α, and 54% of donors based on IL-2 production (**Figure 1A-C**, **Table I S1**). Overall, 92% of donors showed a specific CD4+ T cell response to at least one SARS-CoV-2 protein based on production of at least one cytokine, where a positive response was defined as a 10% increase over control stimulated samples (**Table SI**). No responses were detected to OC43 spike protein by ICC in the same donors (**Figure S1B**), whereas 100% of donors produced cytokines in response to PMA/ionomycin (**Figure S1B**). We did not observe cytokine production in CD8+ T cells from any of the donors, albeit CD8+ T cells from all donors responded to influenza A/PR8/34 virus (PR8) as well as to PMA/ionomycin (data not shown). This is likely because CD8+ T cells respond poorly to whole protein antigens. Taken together, our data show that the vast majority of SARS-CoV-2 convalescent individuals have recall CD4+ responses to SARS-CoV-2 S or N proteins at 4-10 weeks after initial symptoms, with IL-2 and TNF-α predominating over IFN-γ.

**Figure 1.**
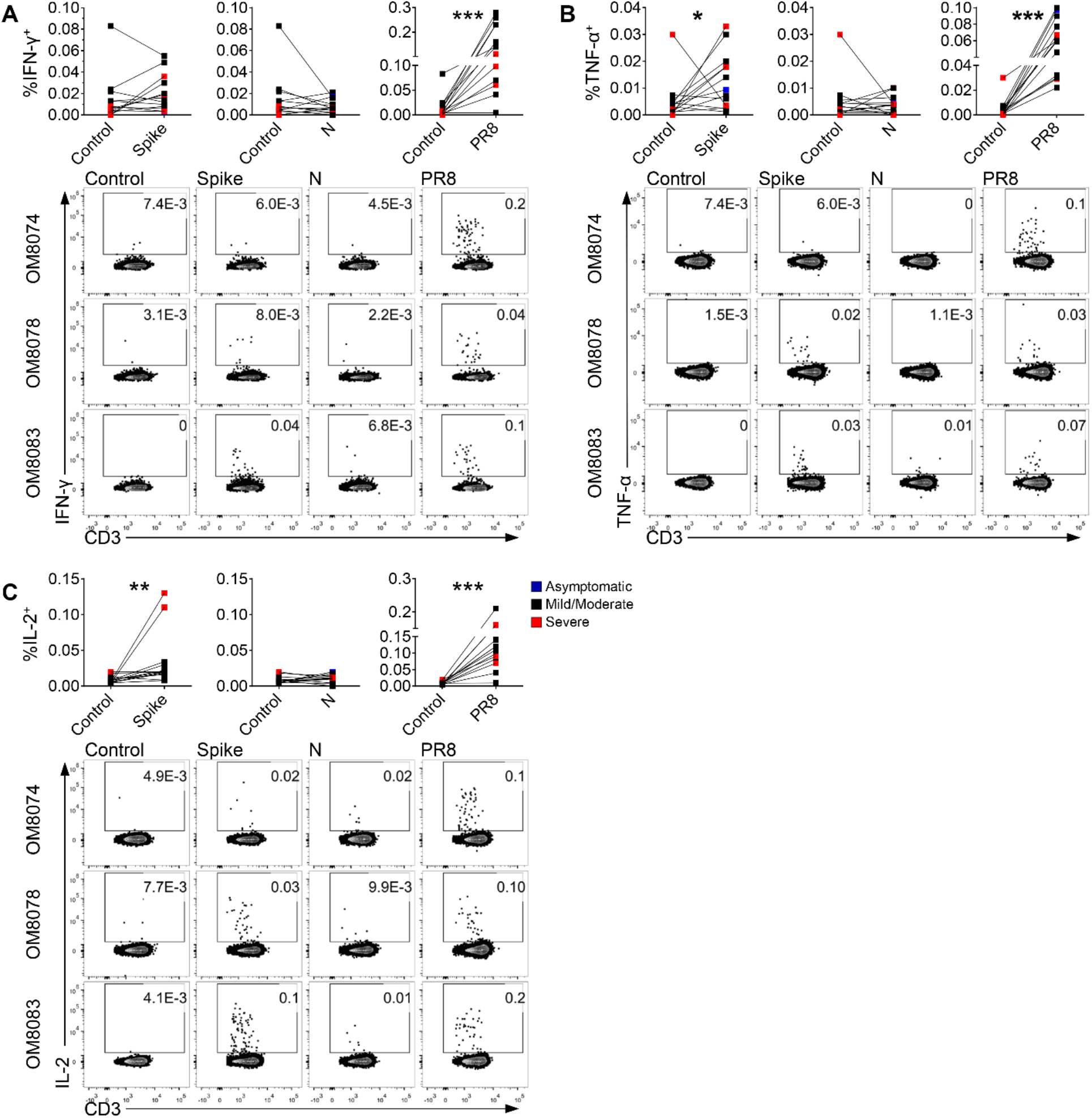
Intracellular cytokine responses to Spike and N proteins in convalescent COVID-19 subjects by flow cytometry. Cytokine production by SARS-CoV-2-specific CD4+ T cells after 18h of incubation with S, N or Influenza A PR8. Graphs and representative flow cytometry plots show the frequency of CD4+ T cells expressing: **(A)** IFN-γ (n=13), **(B)** TNF-α (n=12) and **(C)** IL-2 (n=13). One donor exhibited high background TNF-α+ CD4+ T cells and was determined to be an outlier by the Grubb’s test. Although this data point is shown in all panels, it was excluded from statistical analysis of TNF-α+ CD4+ T cells. Pair-wise comparisons were made in **(A)-(C)** by two-tailed Wilcoxon test. *p<0.05, **p<0.01, ***p<0.001.

### Comparison of the CD4+ T cell response to SARS-CoV-2 versus Influenza A virus by multiparameter flow cytometry

As most adults are expected to have memory T cells specific for seasonal influenza virus, we compared recall responses to influenza A/PR8/34 H1N1 virus (PR8) for all donors. 92% of SARS-CoV-2 convalescent patient samples showed strong CD4+ recall responses to PR8 stimulation based on IFN-γ producing T cells and the frequency of these responding cells was substantially higher than responses to S and N proteins (**Figure 1A**). This was not due to insufficient S protein, as increasing the dose from 1 to 5µg per ml did not increase the frequency of responses (**Figure S2A**). We also obtained similar responses using trivalent inactivated seasonal influenza vaccine (TIV), which contains only influenza proteins (**Figure S2B**). Thus, the weaker response to SARS-CoV-2 S protein compared to influenza proteins is unlikely due to the use of live influenza virus versus recombinant SARS-CoV-2 proteins, albeit it could be impacted by an incomplete set of SARS-CoV-2 epitopes covered by including only 2 of the SARS-CoV-2 proteins. Some human T cell studies use costimulation with anti-CD49d and anti-CD28 to increase the sensitivity of detection with ICC (32), however we found no difference in the frequency of response to S with or without additional costimulation (**Figure S2C**). We also repeated the assays 3 times for 2 of the donors and obtained a similar frequency of responding T cells each time (**Figure S2D**). In contrast to the results with SARS-CoV-2 convalescent patients, PBMC collected from healthy donors in early March 2020 did not show detectable T cell responses to S, N or OC43 S whole proteins, albeit all the healthy donors responded to PR8 (**Figure S3**). As will be discussed below, the lack of detection of cross-reactivity in healthy donors may reflect the relative insensitivity of the ICC assay compared to other methods of detecting cross-reactivity.

Analysis of T cell production of multiple cytokines showed that 80% of S-specific CD4+ T cells produced only 1 cytokine. Influenza-specific CD4+ T cell responses were more multifunctional, with 8.7% of PR8-specific CD4+ T cells as compared to 3.4% of S-specific T cells producing all 3 cytokines (**Figure 2A**). We also noted that the ratio of IFN-γ to TNF-α producing cells was significantly higher among the PR8-specific CD4+ T cells than the S-specific CD4+ T cells (**Figure 2B**) and this was independent of disease severity.

**Figure 2.**
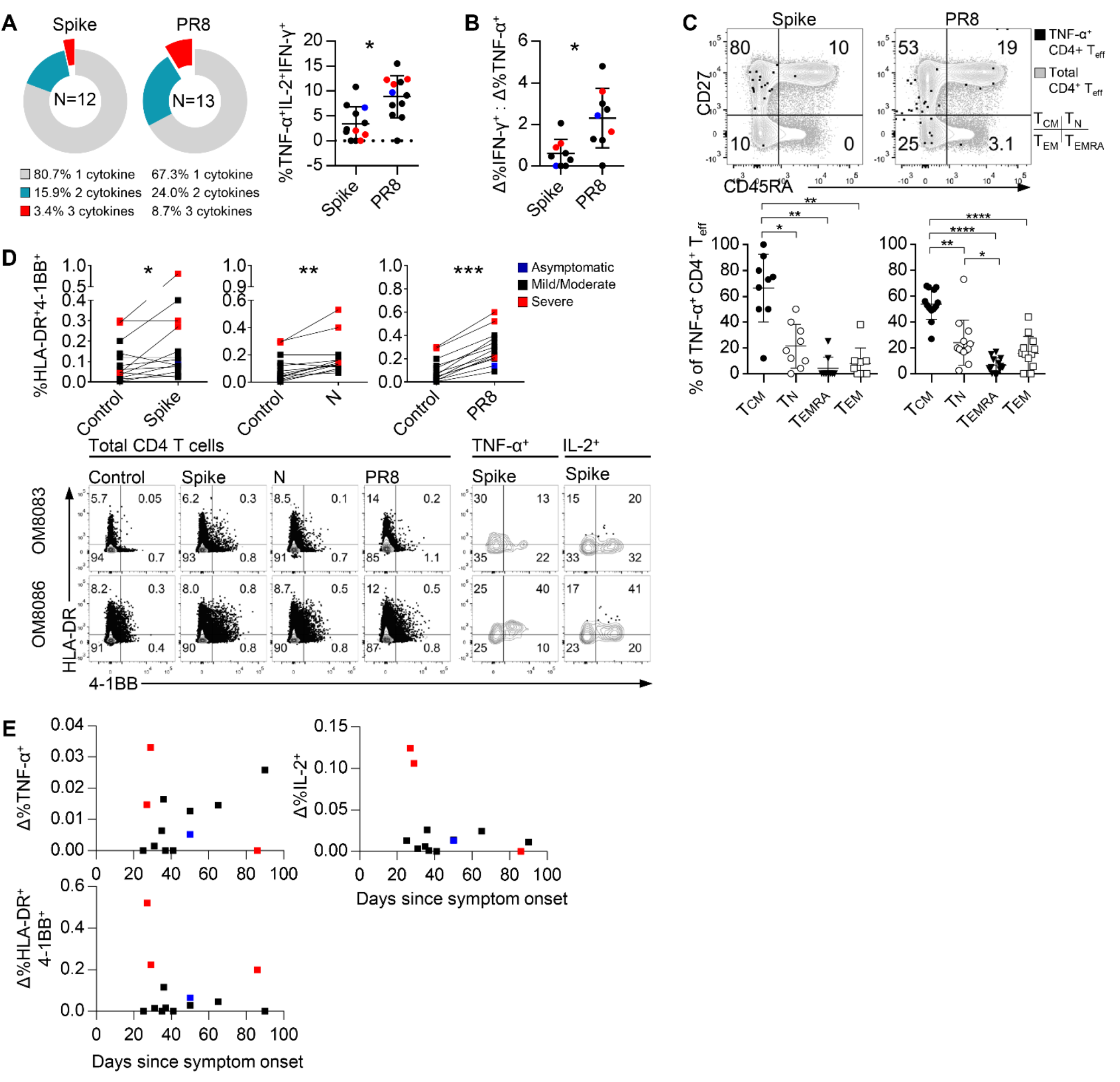
Comparison of CD4+ T cell responses to SARS-CoV-2 or Influenza A virus. **(A)** Frequency of cells expressing IFN-γ, TNF-α and/or IL-2 as a proportion of total cytokine producing cells. **(B)** Ratio of the %IFN-γ+:TNF-α+ CD4+ T cells in donors producing both cytokines (n=9). **(C)** Representative flow cytometry plot of CD27 and CD45RA expression by total CD4+ T cells and TNF-α+ CD4+ T cells (n=9). The distribution of memory subsets of TNF-α+ CD4+ T cells is shown for the donors with a TNF-α response. Graphs show mean±SD. **(D)** Graphs show the %CD4+ T cells co-expressing HLA-DR and 4-1BB. Representative flow cytometry plots show the expression of HLA-DR and 4-1BB by total CD4+ T cells, amd TNF-α+ and IL-2+ CD4+ T cells after stimulation with Spike. **(E)** The frequency of TNF-α+, IFN-γ+ and HLA-DR+4-1BB+ CD4+ T cells versus days since symptom onset. Pair-wise comparisons were made in **(A), (B)** and **(D)** by two-tailed Wilcoxon test and in **(C)** by one-way ANOVA with Holm-Sidak’s multiple comparisons test. *p<0.05, **p<0.01, ****p<0.0001.

Analysis of CD27 and CD45RA expression on the S-specific and PR8-specific TNF-α-producing CD4+ T cells indicated that the responding T cells were predominantly central memory T cells (**Figure 2C**). The activation markers HLA-DR and 4-1BB are frequently used to determine specific recall responses. Based on these markers, 100% of donors responded to influenza PR8, whereas 69% responded to Spike and 85% to N. Examination of HLA-DR/4-1BB double positive cells for cytokine production showed some discordance between activation markers and cytokine producing cells (**Figure 2D**), with neither approach identifying 100% of the responding CD4+ T cells.

Although there appeared to be a trend towards higher responses in donors with severe illness in the first 4 weeks, using either activation markers and/or production of cytokines as a measure of response, differences in ICC response based on disease severity were not significant (**Figure 2E**). Taken together, our data show that SARS-CoV-2-specific T cells from patients in the early convalescent phase are largely of the Tcm phenotype and respond to S and N proteins with a higher ratio of TNF-α:IFN-γ producing cells compared to the response to influenza virus, which shows a more typical anti-viral IFN-γ dominant response.

### Recall responses of SARS-CoV-2 convalescent PBMC based on cytokine secretion

To further analyze cytokine production during recall responses to SARS-CoV-2 we collected supernatants from *ex vivo* PBMC 48hrs post-stimulation with S, N or influenza PR8 by multiplex bead array analysis of 13 cytokines (**Figures 3, S4**). 92% of SARS-CoV-2 convalescent donors showed IFN-γ production in response to SARS-CoV-2 S, whereas 100% showed IFN-γ responses to SARS-CoV-2 N and PR8, albeit the median level of IFN-γ in response PR8 was higher than that observed in response to SARS-CoV-2 N or S (**Figure 3A, S4A**). 100% of patient PBMC produced specific TNF-α responses in response to N, whereas 50% produced TNF-α in response to Spike and 92% in response to PR8. N-specific responses showed substantially higher TNF-α responses than S or PR8-specific responses (**Figure 3B, S4A**). IL-2 was produced in response to S or PR8, but not in response to N-stimulation, whereas IL-10 was produced in all cases, albeit the highest amount of IL-10 was observed in the N-stimulated cultures (**Figure 3 C, D, S4A**). IL-13 was produced in response to S and PR8, but not N, whereas IL-6 was only observed with S and N restimulation and not with PR8 (**Figure 3 E, F**). Similar to our findings with ICC, the ratio of IFN-γ to TNF-α or IL-10 was highest in cultures stimulated with Influenza A virus (**Figure 3G**). We did not detect IL-4, 5 or 17F in any of the cultures (data not shown). IL-9 and 17A were detected in some cultures but did not show consistent increases with S or N stimulation, whereas IL-22 was produced in response to S stimulation for some donors (**Figure S4B**). Healthy donor PBMC produced IL-6, 10, IFN-γ and TNF-α in response to N but not S proteins (**Figure S4C**). Stimulation of SARS-CoV-2 convalescent PBMC as well as healthy donor PBMC with OC43 S protein resulted in induction of IFN-γ, TNF-α, IL-10 and IL-6, but not IL-2 (**Figure S5A, B**).

**Figure 3.**
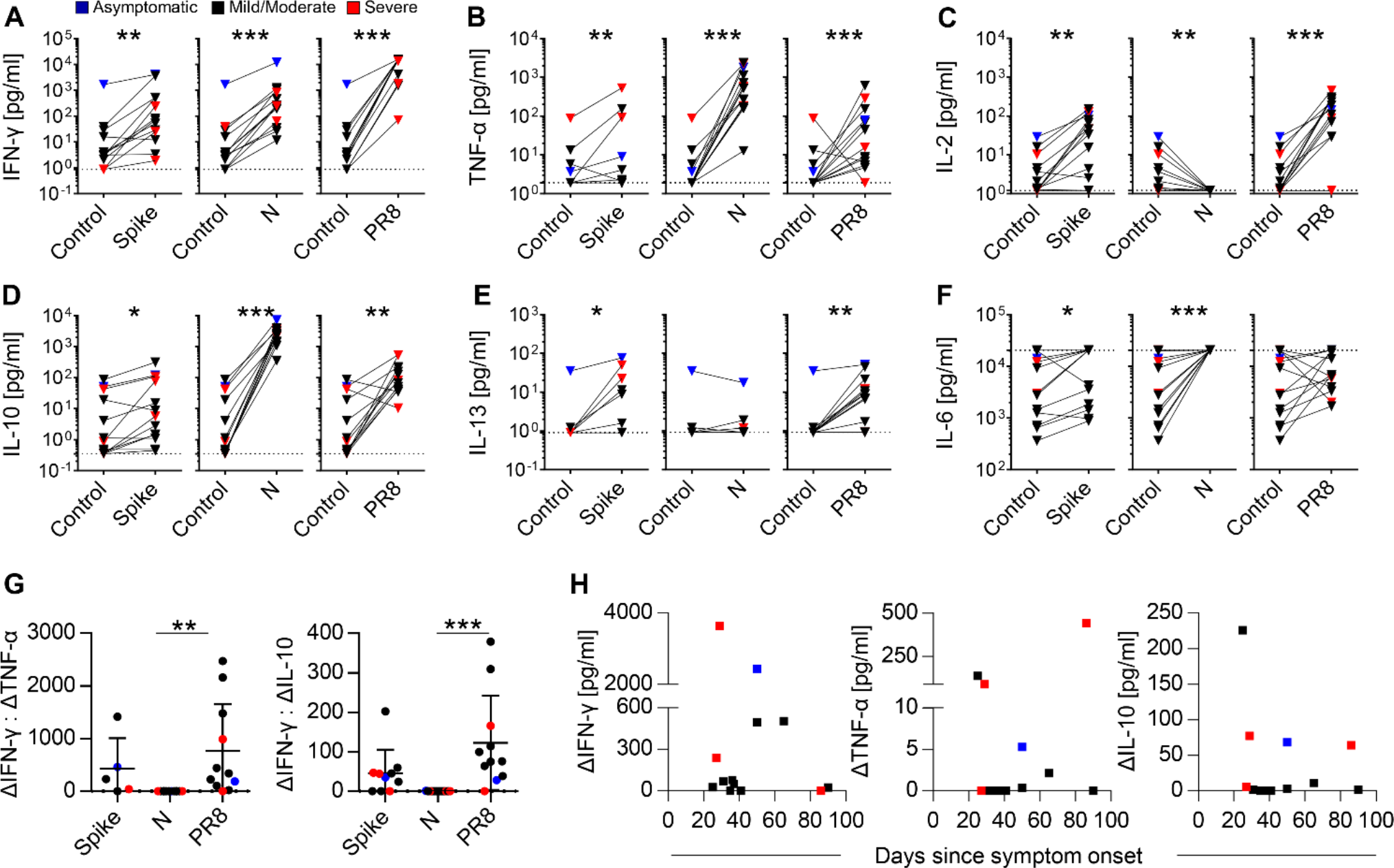
Recall responses of SARS-CoV-2 convalescent PBMC based on cytokine secretion. Cytokines in cell culture supernatants after 48h stimulation with S, N or PR8 as quantified by the multiplex cytokine bead assay (n=13). Graphs show **(A)** IFN-γ, **(B)** TNF-α, **(C)** IL-2, **(D)** IL-10, **(E)** IL-13, and **(F)** IL-6. **(G)** Ratio of IFN-γ: TNF-α and IFN-γ:IL-10 in cell culture supernatants (Spike n=6, N n=11, PR8 n=11). Graphs show mean ± SD. **(H)** The levels of IFN-γ, TNF-α, and IL-10 versus days since symptom onset. OM8099 exhibited high background TNF-α and was determined to be an outlier by the Grubb’s test. Although this data point is shown in **(B)**, it was excluded from statistical analysis of TNF-α responses. Pair-wise comparisons were made by two tailed Wilcoxon test for **(A)-(F)**. Nonparametric Dunn’s multiple comparisons test was performed for **(G)**. Graphs show mean±SD. *p<0.05, **p<0.01, ***p<0. 001.

Overall, the multiplex cytokine assays show a predominant Th1 profile based on restimulation with SARS-CoV-2 or seasonal hCoV-OC43 S protein, as well as reactivity of healthy donor PBMC to SARS-CoV-2 N and OC43 S protein. SARS-CoV-2 convalescent patients’ PBMC showed a lower IFN-γ to TNF-α or IFN-γ to IL-10 in response to SARS-CoV-2 proteins compared to influenza virus restimulation. Particularly striking was the consistent and high-level production of IL-10 in response to N in all SARS-CoV-2 convalescent PBMC tested.

### Peripheral T follicular helper as well as T effector responses to SARS-CoV-2 antigens correlate with serum antibodies and neutralization titers

T follicular helper (Tfh) responses are important for the generation of long-lived antibody responses (33). Although fully differentiated Tfh are normally found in the lymphoid organs, their peripheral blood precursors, pTfh, can be detected in the blood based on expression of CCR7 and CXCR5 (34, 35). Here we used expression of IL-2 by ICC combined with CCR7 and CXCR5 expression to detect antigen-specific pTfh cells in SARS-CoV-2 convalescent patient PBMC following restimulation with S, N or PR8 (**Figure 4A**). 46% of samples showed S-specific IL-2 producing pTfh, 54% N-specific and 100% of PBMC samples showed PR8-specific pTfh. For samples collected during the first 4 weeks post-symptoms, pTfh responses to S were higher in severe compare to mild cases (**Figure 4A, B**). We also compared pTfh responses of PBMC from the SARS-CoV-2 convalescent patients with IgG and IgA responses to N and RBD based on serum ELISA (36) and neutralization data (**Figure 4C,D**). There was a positive correlation between the frequency of IL-2+pTfh and N-specific IgG (R=0.57, p<0.05). The correlation between IL-2+pTfh and RBD-specific IgG showed a similar positive trend (R=0.29) but did not reach statistical significance. Similarly, no significant correlation with S-specific IgG was observed (data not shown). There was a significant positive correlation between the pTfh response to RBD-specific IgA (R=0.66, p< 0.05). There was also a strong correlation between WT SARS-CoV-2 IC_50_ neutralization (modified PRNT assay) by patient sera and the pTfh response (R=0.84, p<0.001). Similar correlations were obtained by calculating the area under the curve (AUC) in a surrogate neutralization ELISA with patient sera, human ACE2 and immobilized S-RBD (R=-0.75, p<0.01) (**Figure 4E**).

**Figure 4.**
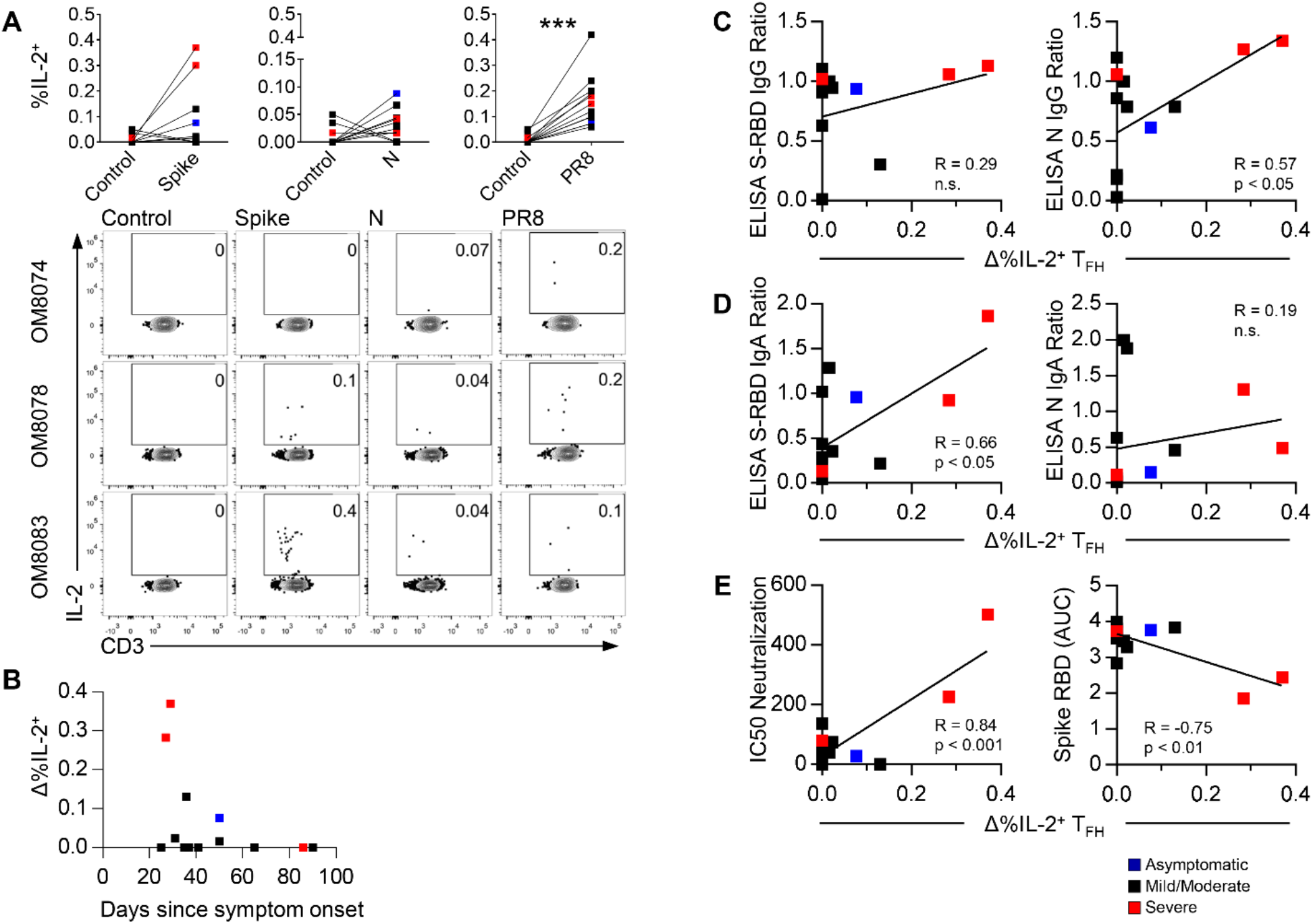
pTfh responses to SARS-CoV-2 antigens. **(A)** Graphs and representative flow cytometry plots show %IL-2+ pTfh cells in response to Spike, N and PR8 after 18h of stimulation (n=13). **(B)** %IL-2+ pTfh versus days since symptom onset. **(C)** Correlation between S-RBD or N serum IgG and %IL-2+ pTfh. **(D)** Correlation between S-RBD or N serum IgA and %IL-2+ pTfh. **(E)** Correlation between viral neutralization titres and %IL-2+ pTfh or between S-RBD IgG AUC in a surrogate neutralization ELISA with human ACE2 and %IL-2+ pTfh. Serum antibody titres were normalized to a positive control well. Pair-wise comparisons were made by two-tailed Wilcoxon test for (A). Correlation analysis for **(C)-(E)** was performed by Pearson’s correlation. ***p<0.001.

Similarly, there was a positive correlation between the frequency of IL-2+ CD4+ T cells and N-specific IgG (R=0.61, p<0.05), and a positive trend between IL-2+ CD4+ T cells and RBD-specific IgG (R=0.36), albeit not significant (**Figure 5A**). There was also a positive correlation between IL-2+ CD4+ and RBD-specific IgA (R=0.60, p<0.05) (**Figure 5B**). A strong positive correlation was also found between virus neutralization and IL-2+ CD4+ T cells (R=0.82, p<0.001), and between S-RBD IgG AUC and IL-2+ CD4+ T cells. (**Figure 5C**). There was also a positive correlation between virus neutralization and disease severity (**Figure 5D**).

**Figure 5.**
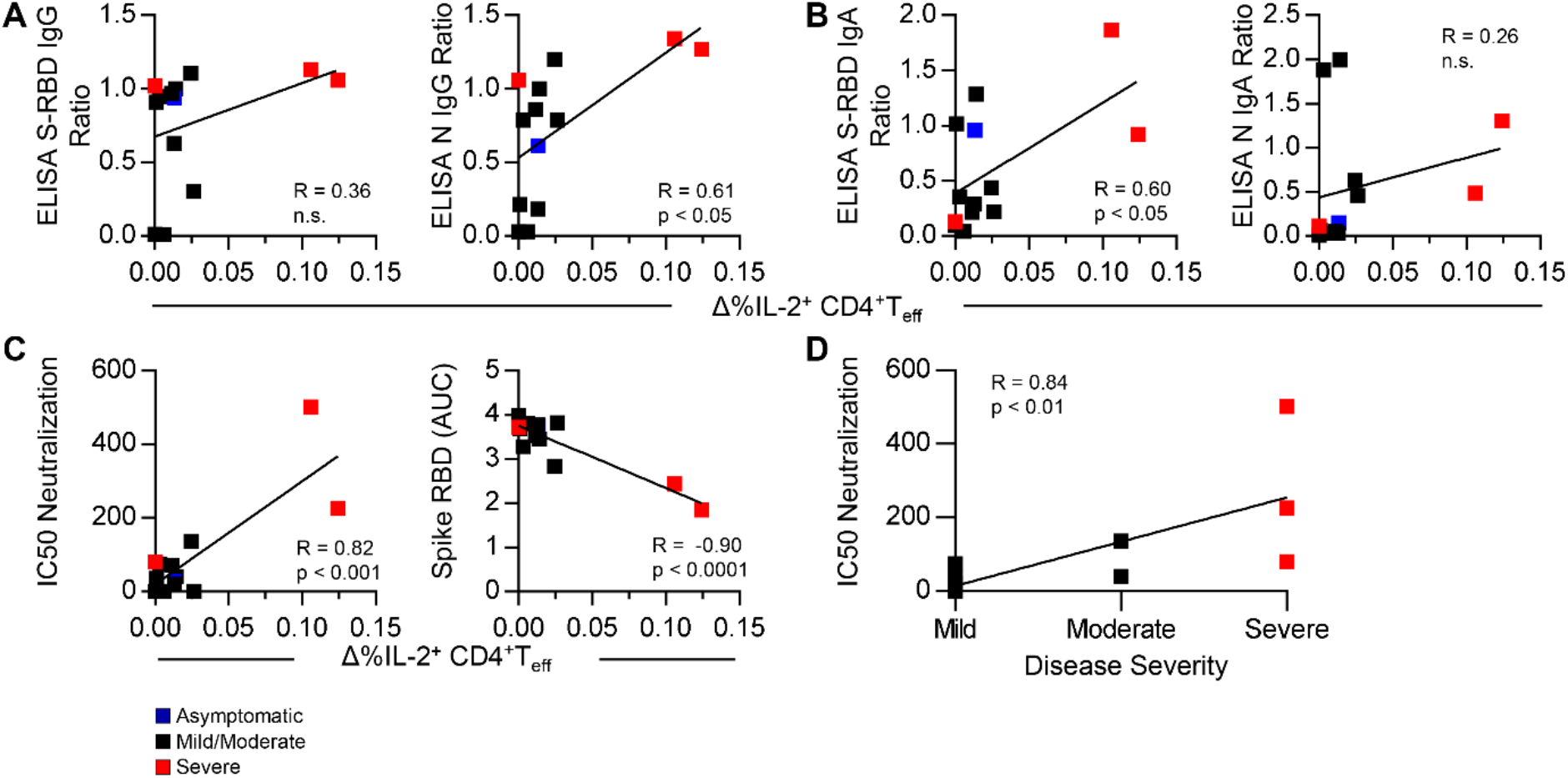
Correlation analysis between CD4+ T cell responses, serum antibodies and virus neutralization. Correlation analysis was performed between **(A)** S-RBD or N serum IgG and %IL-2+ CD4+ T cells, **(B)** between S-RBD or N serum IgA and %IL-2+ CD4+ T cells, **(C)** between virus neutralization titres or S-RBD IgG AUC and %IL-2+ CD4+ T cells, and **(D)** between virus neutralization titres and disease severity. Serum antibody titres were normalized to a positive control well. Correlation analysis was performed by Pearson correlation in (A)-(C), and by Spearman correlation in **(D)**.

Thus, pTfh responses can be detected in 6 out of 13 SARS-CoV-2 convalescent PBMC responding to S protein, 7/13 in response to N whereas 13/13 showed a Tfh response to influenza A virus. Both pTfh and T effector responses correlated strongly with the neutralization titers observed in the same donors, with the highest neutralization activity correlating with disease severity.

### CD4+ and CD8+ T cell proliferative responses to peptide pools

Much of the published work on SARS-CoV-2 specific T cells has focused on peptide pools and these are more effective in inducing CD8+ recall responses than intact proteins. Therefore, we used peptide pools encompassing the RBD, transmembrane (TM) and cytoplasmic regions of S as well as N, Envelope (E) and Matrix (M) to stimulate PBMC from the same patient samples used for ICC. As T cell proliferation to virus antigens has previously been associated with ability to control the virus (37, 38), we labelled PBMC CFSE and assessed the proliferation of the T cells after 7 days by flow cytometry in response to each peptide pool (**Figure 6A**, with gating strategy shown in **Figure S6**). Proliferation responses of total T cells to at least one antigen was observed in 12/13 donors (**Table S2**). There was considerable variability between donors. Generally, donors who made strong proliferative responses had strong responses to all antigens tested. However, antigen-specific proliferation did not correlate significantly with disease severity (**Figure 6B, C, D**). We also examined PBMC from a SARS-CoV-1 patient taken 17 years post-illness and observed modest reactivity to the N peptide pool (**Figure 6B, C**). We next broke down responses into CD4+ and CD8+ T cell responses for each peptide pool. For most subjects and antigens, CD4+ T cell proliferative responses were substantially higher than CD8+ T cell responses, independent of disease severity (**Figure 6E**).

**Figure 6.**
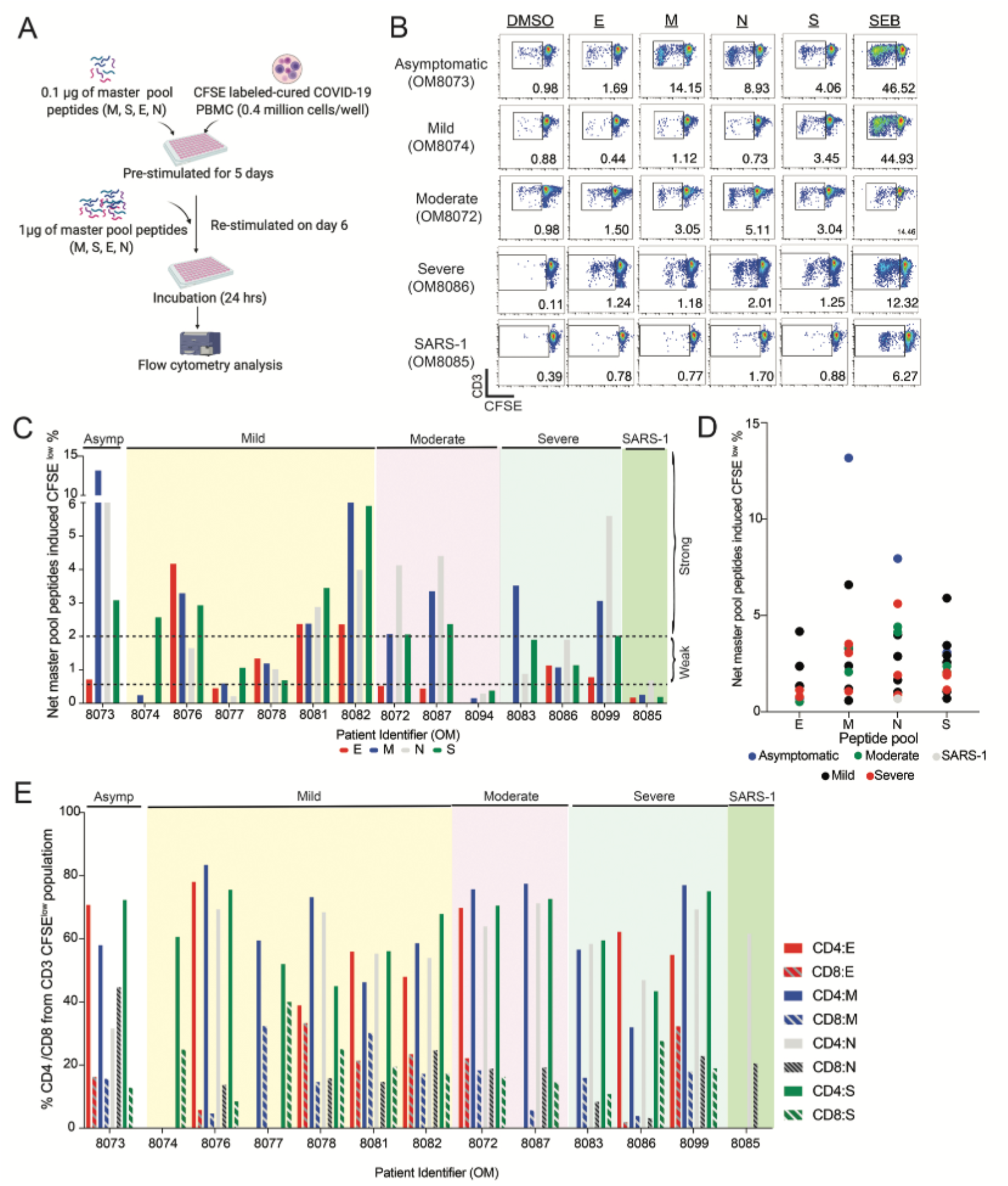
T cells proliferation responses induced by master pool peptides (E, M, N, S) in convalescent COVID-19 patients. (**A)** T cell proliferation assay setup. PBMCs were pre-labeled with CFSE, pre-stimulated with 0.1μg/ml of master pool peptides for 5 days, then restimulated with 1μg of master pool peptides on day 6 for 24 hours. **(B)** Representative flow cytometry plots of CFSE fluorescence by CD3+ T cells. **(C)** Net master pool peptides induced T cell proliferative responses from convalescent asymptomatic (Asymp, n=1), mild (n=6), moderate (n=3), severe (n=3) and SARS-1 (n=1) patients. Net CFSE^low^ percentages were calculated by subtracting the DMSO stimulated percentages from the master pool peptides. The horizontal dashed line at 0.5% and 2.0% indicates weak and strong positive responses, respectively. **(D)** Comparison of T cell proliferative responses from asymptomatic (n=1), mild (n=6), moderate (n=2), severe (n=3) and SARS-1 (n=1) convalescent patients against master pool peptides (E, M, N, and S). **(E)** The frequency of CD4+ and CD8+ T cells within CFSE^low^ CD3+ T cells in each patient.

Although CD8+ cytotoxic T cells are classically associated with virus-infected cell killing, CD4+ granzyme+ cytotoxic T cells can be a significant part of the human anti-viral T cell responses (37, 39-41). Therefore, we also assessed IFN-γ and granzyme B levels by flow cytometry in the CFSE^low^ responding CD4+ and CD8+ T cells (**Figure 7A, B**). Of note, the samples from subjects with severe and moderate disease tended to have a higher proportion of CD4+ IFN-γ/granzyme B co-producing T cells than mild, however this was not universally the case, as we also saw a high proportion of IFN-γ/granzyme B expressing T cells in the asymptomatic donor.

**Figure 7.**
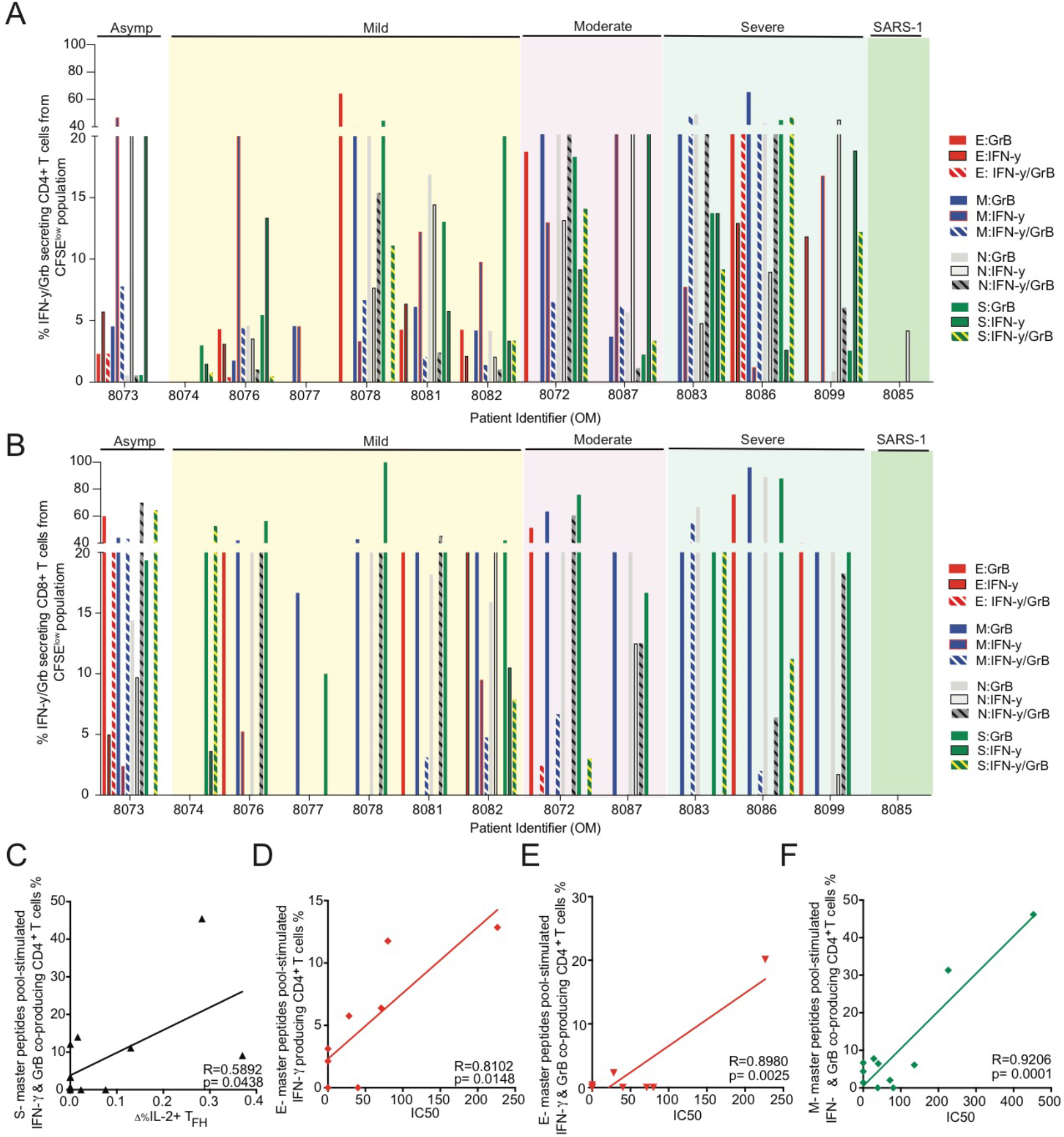
IFN-γ/Granzyme B producing proliferating T cells. The percentage of IFN-γ/Granzyme B co-producing **(A)** CD4+ or **(B)** CD8+ from CD3+ CFSE^low^ T cells from convalescent asymptomatic (n=1), mild (n=6), moderate (n=2), severe (n=3) and SARS-1 (n=1) patients. **(C)** Correlation analysis between S-master peptide pool stimulated IFN-γ and Granzyme B co-producing CD4+ T cells and %IL-2+ pTfh cells in response to Spike. Correlation analysis between virus neutralization titres (IC_50_) and **(D)** E-master peptides pool stimulated IFN-γ producing CD4+ T cells, or **(E)** E-master peptides pool stimulated IFN-γ and Granzyme B co-producing CD4+ T cells or (F) M-master peptides pool stimulated IFN-γ and Granzyme B co-producing CD4+ T cells. Pearson’s correlation test (n=12, SARS-1 patient excluded). Asymp: asymptomatic.

The frequency of proliferating IFN-γ/granzyme B co-producing T cells in response to S peptide pools correlated with the frequency of IL-2 producing pTfh in response to intact Spike (**Figure 7C**) as well as with virus neutralization titers (**Figure 7D**). In addition, proliferating IFN-γ/granzyme B producing cells in response to E or M peptide pools correlated with serum neutralization titers (**Figure 7 E, F**). Thus, a strong CD4+ response overall, whether based on analysis of whole protein or peptide stimulation, correlates with strong neutralization responses.

## Discussion

In this study, we have conducted a systematic examination of T cell recall responses in PBMC taken in the early convalescent phase (4-12 weeks post-symptoms) of COVID-19 in response to SARS-CoV-2 recombinant proteins as well as to peptide arrays. The use of recombinant proteins is relevant because it allows us to assess the response in the context of antigen presentation, and the use of the fully glycosylated Spike trimer is important in mimicking the form of antigen that is presented by intact virus. A T cell response was detected in all SARS-CoV-2 convalescent patients, with 92% responding based on ICC responses to recombinant SARS-CoV-2 proteins and 100% responding based on multiplex cytokine assays. CD8+ T cell responses were not detected in response to whole protein restimulation by ICC but were identified in 12 out of 13 patients based on proliferation in response to restimulation with peptide pools encompassing N, E, M or S, albeit with varying frequencies. While several other studies have identified Th1 responses in response to SARS-CoV-2 peptide or protein stimulation (11, 23, 25, 27, 29), our comparison of recall responses to influenza within the same donors highlights some key differences in SARS-CoV-2 versus influenza-specific T cell responses.

Intracellular cytokine staining revealed that SARS-CoV-2-specific CD4+ recall responses exhibit a hierarchy of IL-2>TNF-α>IFN-γ, whereas influenza A virus-specific T cells show IFN-γ >IL-2> TNF-α based on frequency of cytokine producing cells. This altered Th1 profile in SARS-CoV-2-specific T cells could contribute to increased inflammation with poorer viral control compared to influenza virus-specific T cells. It was possible that these differences are due to the use of whole influenza A virus compared to recombinant proteins. However, control experiments showed indistinguishable frequencies of responding CD4+ T cells producing IFN-γ, TNF-α and IL-2 whether samples were restimulated with live influenza virus or with TIV, the inactivated influenza vaccine which is dominated by the hemagglutinin protein. It is unlikely that the altered cytokine response in response to SARS-CoV-2 antigens is driven by disease severity, as mild and severe patients were distributed throughout the plots showing this altered ratio (Fig. 2B, 3G). On the other hand, the lower frequency of multifunctional cells in the SARS-CoV-2 specific as compared to influenza A-specific CD4+ recall responses seems to be heavily weighted by the severe cases (Fig. 2B) and could reflect COVID-19-specific exhaustion, as has been suggested by other studies based on activation/exhaustion markers (42, 43). The influenza-specific recall responses we observed are likely due to a lifetime of seasonal exposure and/or vaccination, whereas the SARS-CoV-2-specific responses are more recent. However, it is unlikely that the time since exposure is driving the altered cytokine profile we observe, as the cytokine profile observed in recall responses generally reflects the epigenetic profile imprinted during priming (44).

Multiplex analysis of cytokines in supernatants of PBMC following SARS-CoV-2 antigen stimulation also revealed lower IFN-γ to TNF-α ratios of SARS-CoV-2 compared to influenza-specific responses as well as higher levels of IL-10 and IL-6. The ICC flow cytometry assay allows one to clearly identify the source of the cytokines as CD4+ T cells, whereas the multiplex cytokine assay reflects total amount of secreted cytokine and can also reflect cytokines secreted from other cells, such as monocytes or NK cells, in response to the activated T cells. IL-10 can be produced by both T cells and antigen presenting cells, whereas IL-6 is likely coming from monocytes responding to the activated T cells in the PBMC culture. Particularly striking in our study was the high level of IL-10 detected in the supernatants of N-stimulated cultures, which could contribute to impaired antigen presentation and immunosuppression (45). Further investigation is required to determine whether N-specific responses are immunosuppressive, which would have significant implications for vaccine design.

A potential caveat to our findings is that we included only 2 of the SARS-CoV-2 proteins in our cytokine analysis and not the full spectrum of SARS-CoV-2 antigens. However, the cytokine profile we observed in the supernatants of S and N stimulated PBMC is quite similar to that reported by Weiskopf et al for SARS-CoV-2 ARDS patient PBMC collected 3 weeks after ICU admission and stimulated with peptide megapeptide pools covering most of the SARS-CoV-2 proteins (23). There were, however, some differences noted, such as their detection of IL-17A, which we did not detect in our assays.

The cytokine profile we detect in the supernatants of SARS-CoV-2 convalescent PBMC after antigen stimulation is similar to the overall cytokine profile reported at the acute phase of infection, including high levels of IL-6, IL-10 and TNF-α (46, 47). This is consistent with the evidence that memory T cells are imprinted by the acute inflammatory milieu (44). Schultheiss et al.(48) recently analyzed total PBMC from SARS-CoV-2 active and early convalescent patients and also noted that total CD4+ T cells showed an altered non-classical Th1 profile, similar to what we observe here with antigen-specific T cell responses. They also noted Th17 responses, which were not consistently observed in the antigen-specific T cells in our cohort.

Of note, we observed a disconnect between ICC responses and analysis of T cell responses to S and N based on activation markers. This was not unique to SARS-CoV-2, however, as we observed a similar disconnect with influenza A-specific T cell cytokine response and activation markers (data not shown). It is possible that some antigen-specific T cells are not making cytokines in the time frame analyzed or that some of these activation markers are induced on memory T cells by bystander effects (cytokines). We suggest, that functional readouts based on cytokines may be more relevant to understanding protective immunity to SARS-CoV-2 than use of activation markers.

Peripheral T follicular helper responses (IL-2+CCR7+CXCR5+) were detected in 62% of PBMC after S or N stimulation, and strongly correlated with virus neutralization activity of sera based on neutralization of SARS-CoV-2 as well as a surrogate neutralization ELISA for binding to RBD. The strong correlation with IgA might reflect the recently reported role for IgA in SARS-CoV-2 neutralization (49). Our findings are similar to those of Ni et al. (11) who showed a correlation between total N-specific T cells measured by ELISpot with neutralizing antibody titers. Of note, total T effector responses to N and S as well as IFN-γ or IFN-γ and granzyme B responses to E and M peptide pools also correlated with virus neutralization, suggesting that a strong CD4+ T cell response in general correlated with effective virus neutralization, whether we used peptide pools or intact antigens for these recall assays. Of note, 100% of donors showed pTfh responses to influenza virus and the response was generally of higher frequency. Thus, the Tfh response to SARS-CoV-2 in convalescent subjects is weaker than that observed in response to influenza virus restimulation.

Several recent studies have revealed responses of healthy donors to SARS-CoV-2 peptides (21-23, 28, 31). It has also been suggested that prior exposure to seasonal coronaviruses might allow some cross-protective immunity to SARS-CoV-2 (30). Our ICC assays did not reveal responses of SARS-CoV-2 convalescent patients or healthy donors to seasonal OC43 spike protein. However, such responses were detected in supernatants based on the cytokines IFN-γ, TNF, IL-6 and IL-10, but not IL-2. This may reflect the lower sensitivity of the overnight ICC assay, compared to assessment of cytokines in the supernatant at 48hrs. Healthy donors similarly responded to OC43 spike but with approximately 10-fold weaker responses than SARS-CoV-2 convalescent patients suggesting that recent boosting with SARS-CoV-2 might enhance such responses. Healthy donors also responded to SARS-CoV-2 N but not S based on release of IFN-γ, TNF-α and IL-6 in the supernatant. We also detected proliferative responses to the N peptide pool of a SARS-CoV1 patient, 17 years post-illness, similar to results recently reported (21).

Proliferative responses to SARS-CoV-2 peptide pools showed that CD4+ responses predominated over CD8+ T cell responses, which might contribute to the pathophysiology of COVID19. On the other hand, many of these CD4+ T cells co-produced IFN-γ and granzyme B, suggesting cytotoxic potential. As airway epithelial cells, the target of SARS-CoV-2 infection, can express MHC II (50-52), these granzyme B positive cells may be relevant to viral control.

A recent study of early T cell responses to SARS-CoV-2 showed delayed T cell responses, compared to antibody responses in the first two weeks post symptom onset, but with T cell responses increasing at >3 weeks (53). Although we did not do a kinetic analysis, the data on convalescent samples collected at 4-12 weeks post-symptoms, are consistent with a peak response around 4 weeks, and falling off thereafter. These kinetics are similar to what was observed in the recall response to the 2009 influenza virus pandemic, where peripheral blood CD4+ and CD8+ T cell responses to whole H1N1 restimulation peaked at about 3-4 weeks post symptoms and then fell off gradually (54). We did not see a consistent difference between severe and mild cases in terms of magnitude of the T cell response, albeit this may be limited by sample size.

In sum, our study shows robust T cell recall responses in SARS-CoV-2 convalescent subjects at 4-12 weeks post-symptoms. Based on proliferation, ICC or multiplex ELISA, all donors showed SARS-CoV-2-specific T cell responses. By 4 weeks post-SARS-CoV-2 infection, most subjects exhibit a strong CD4+ Th1 recall response, with a less predominant CD8+ T cell response and an altered cytokine profile with more TNF-α and less IFN-γ compared to responses to influenza virus in the same donors. In addition, pTfh responses to SARS-CoV-2 were weaker than that to influenza A virus. SARS-CoV-2 N-specific T cell responses were associated with strong induction of IL-10, suggesting that N protein might contribute to immunosuppression. This could have important implications for vaccine design. Taken together, these results suggest that CD4+ T cell responses are more inflammatory than influenza-specific recall responses and show a weaker Tfh response, potentially contributing to disease. The strong correlation between the N- or S-specific pTfh response or the IFN-γ/granzyme B+ proliferative response and neutralization capacity suggests that these responses should be incorporated into vaccine design and testing.

## Methods

### Human subjects and study approval

Written informed consent was obtained from COVID-19 convalescent and healthy blood donors before leukapheresis or peripheral blood samples were obtained. Individuals with recovered COVID-19 infection that was confirmed by positive nasopharyngeal COVID-19 PCR upon presentation, were leukapheresed after resolution of symptoms through an REB approved protocol (St. Michael's Hospital REB20-044c to MO. Additional healthy donors were recruited at the University of Toronto (REB# 00027673 to THW). All human subjects research was done in compliance with the Declaration of Helsinki.

### Human PBMC isolation

PBMCs were isolated from whole blood of healthy human donors by density centrifugation using Ficoll-Paque PLUS (GE Healthcare). PBMCs were cryopreserved in 10% DMSO in AIM-V media (Gibco) before use.

### Virus and viral antigens

The human codon-optimized cDNA encoding the OC43 spike protein (AAT84354.1) was synthesized by GeneArt. The soluble OC43 spike construct includes residues 15-1295, followed by a T4 fibritin trimerization motif, a TEV cleavage site, and a 6xHis-tag. The 20 amino acid human cystatin secretion signal was added N-terminal to the spike sequence. To stabilize the pre-fusion state of the OC43 spike trimer, residues 1070-1071 (AL) were mutated to two proline residues (PP) as described for other spike proteins (55). The human codon-optimized cDNA encoding the SARS-CoV-2 spike protein (YP_009724390) was synthesized by GenScript. The soluble spike trimer construct includes residues 1-1211, followed by a T4 fibritin trimerization motif, a 6xHis-tag and an AviTag biotinylation motif (56). Residues 682-685 (RRAR) were mutated to SSAS to remove the furin cleavage site on the SARS-CoV-2 spike protein. Residues 986-987 (KV) were mutated to two proline residues (PP) to stabilize the pre-fusion form.

The spike proteins were cloned into a piggyBac-based inducible expression vector PB-T-PAF. Inducible stable cell lines were generated in Freestyle 293-F cells (Thermofisher) as previously described (36, 57). For the OC43 spike protein, the stable cells were grown as an adherent culture in DMEM/F12 medium supplemented with 3% (v/v) FBS. For the SARS-CoV-2 spike protein, the stable cells were grown in suspension culture in Freestyle 293 expression medium (Thermofisher). Protein expression was induced by the addition of 1 µg/mL doxycycline. The secreted proteins were purified from the tissue culture medium using Ni-NTA resin. The proteins were further purified by size-exclusion chromatography using a Superose 6 Increase column (GE healthcare). The quality of the purified spike protein trimers was assessed using negative stain electron microscopy.

Nucleocapsid^1-419^ (N) expressed as a N-terminally tagged HIS-GST-TEV fusion was purified from bacteria and kindly provided by Frank Sicheri, Mt. Sinai Hospital, as described in (17).

Endotoxin levels were measured in S and N proteins using the Toxin Sensor Chromogenic LAL Endotoxin Assay Kit, from GenScript/VWR, Cat # L00350C. Final concentrations of LPS was <1 EU per well (0.18 for S, 0.43 for N). Influenza virus strain A/Peurto Rico/8/1934 (PR8) was grown in embryonated chicken eggs and tissue culture infectious dose determined by infection of MDCK cells (58).

15-mer peptides overlapped by 11 amino acids spanning most of full protein sequence of N, membrane (M), envelope (Env), and RBD/TM/cytoplasmic domains of S protein of SARS-CoV-2 were synthesized (GeneScript). To stimulate PBMC, N-master peptide pool with102 peptides, Env-master peptide pool with 12 peptides, M-master peptide pool with 49 peptides and S-master peptide pool with 49 peptides were used in the study.

### T cell stimulation assay

For all stimulation assays, cryopreserved PBMCs were thawed at 37°C, washed twice with PBS and cultured in complete media (RPMI 1640 supplemented with 10% FBS, 2-ME, sodium pyruvate, penicillin, streptomycin and non-essential amino acids (Gibco)) at 37°C with 5% CO_2_. 2×10^6^ PBMCs were plated per well in 96-well round bottom plates for 18h with 1 µg/ml S, 1 µg/ml N, 3 µg/ml OC43 S or 100 HAU/ml live PR8. PBMCs were cultured with 1 µg/ml BSA (Sigma-Aldrich) as a negative control. GolgiStop (BD Biosciences) containing monensin and GolgiPlug (BD Biosciences) containing brefeldin A was added in the last 6h of the culture. As a positive control, 50 ng/ml PMA (Sigma-Aldrich), 1 µg/ml ionomycin (Sigma-Aldrich), GolgiStop and GolgiPlug were added to PBMCs cultured with complete media in the last 6h of culture.

To assess T cell recall responses to live PR8 compared to TIV (FLUZONE® High-Dose), cells were either cultured with complete media, 100 HAU/ml PR8 or 1 µg/ml TIV for 18h. To determine whether the addition of agonistic co-stimulatory antibodies increased the sensitivity of detection of ICC by flow cytometry, PBMCs were stimulated with 1ug/ml S or 1 µg/ml BSA, either with or without 2 µg/ml anti-CD28 and 2 µg/ml anti-CD49d (BD Biosciences) for 18h. GolgiStop and GolgiPlug were added in the last 6h of these cultures.

### Intracellular cytokine staining

After culture, PBMCs were washed with PBS containing 2% FBS (FACS buffer). Cells were first stained with anti-human CCR7 at 37°C for 10 min, followed by staining with Fixable Viability Dye eFluor™ 506 (eBiosciences) to discern viable cells, and with anti-human CD3, CD4, CD27, CD45RA, CXCR5, 4-1BB and HLA-DR for 20 min at 4°C. Cells were washed twice with FACS buffer, then fixed with BD Cytofix/Cytoperm buffer (BD Biosciences) for 20 min. Following fixation and permeabilization, cells were washed twice with 1X BD Perm/Wash buffer (BD Biosciences) and stained with anti-human IFN-γ, TNF-α, IL-2 and IL-17A for 15 min at 4°C. Antibodies used are as listed in **Table S3**. Samples were washed twice, then resuspended in FACS buffer and acquired on the BD LSRFortessa X-20 flow cytometer using FACSDiva software.

### Multiplex cytokine bead assay

2×10^6^ PBMCs were seeded per well in 96-well round bottom plates with 1 µg/ml S, 1 µg/ml N, 3 µg/ml OC43 S, 1 µg/ml BSA or 100 HAU/ml PR8. Cell culture supernatants were collected after 48h of incubation. Cytokines in the supernatants were measured using the Human Th Cytokine Panel (12-plex) LEGENDplex kit (Biolegend) with capture reagents specific for IL-2, IL-4, IL-5, IL-6, IL-9, IL-10, IL-13, IL-17A, IL-17F, IL-22, IFN-γ and TNF-α. The assay was performed as per the manufacturer’s instructions using a V-bottom plate. Samples were acquired on the BD LSRFortessa X-20 flow cytometer.

### CFSE T cell proliferation assay

PBMC (2 x 10^6^ cells/ml) were pre-labeled with 5µM of carboxyfluorescein diacetate succinimidyl ester (CFSE; Thermo Fisher Scientific) in PBS with 2.5% FBS for 8 minutes in 37°C water bath. Excessive CFSE dye was removed by using 100% FBS and further rinsed with R-10 [RPMI1640, FBS, Pen/Strep (Thermo Fisher Scientific), Glutamax (Thermo Fisher Scientific) and sodium pyruvate (Thermo Fisher Scientific)]. Cells were then resuspended in R-10 [supplemented with IU-IL 2 (BioLegend) and 2-mercaptoethanol (Themor Fisher)] and plated at 0.4x 10^6^ cells per well in a 96-well round-bottomed polystyrene plate at a final volume of 200µ1 These cells were pre-stimulated with 0.1 µg of S, E, N and M master peptide pools or DMSO (negative control) or SEB (positive control) for 5 days. At day 6, cells were re-stimulated with 1µg/ml of master peptide pools and the exocytosis was blocked by the addition of BD GolgiStop and BD GolgiPlug for another 24 hours. At day 7, cells were prepared for flow cytometry staining. LIVE/DEAD™ fixable blue dead cell stain (Thermo Fisher Scientific) was used to determine the viability of cells and then pre-blocked with Fc receptor blocking solution (Human TruStain FcX^TM^; BioLegend) prior to extracellular staining with anti-human CD3 [APC-Cy7: clone SK7 (BD)], anti-human CD4 [BV711: clone SK3 (BD)] and anti-human CD8 [PE: clone HIT8a (BD)]. Cells were then fixed with BD Cytofix and permeabilized with BD Perm/Wash as per the manufacturer’s protocol and stained with anti-human IFNg-[APC: clone 4S.B3 (BD)] and anti-human granzyme B [BV421: clone GB11(BD)]. Samples were acquired on the BD LSRFortessa X-20 flow cytometer. Net peptide pool induced CFSE^low^ responses were calculated as the percentage of CFSE^low^ cells after stimulation with master pool peptides minus the percentage of CFSE^low^ cells after stimulation with DMSO.

### WT SARS-CoV-2 Neutralization Assay

100 μl of Vero E6 cells were seeded into a 96 well plate at 0.3×10^6^ cells/mL and were incubated overnight for attachment. The following day, patient serum was heat inactivated at 56°C for 30 minutes, then serially diluted 8 times, 2-fold downwards starting at 1:10. Equal volumes of SARS-CoV-2 were added to all wells with a final concentration of 100 TCID/well. The plate was incubated for 1h, shaking every 15 minutes. After incubation, all the media from the Vero E6 cells was removed and 50 μl of the SARS-CoV-2/Serum co-culture was used to inoculate the Vero E6 cells. The infection was done for 1h, shaking every 15 minutes. After infection, the inoculum was removed and growth media was added. CPE was tracked over the course of 5 days. Samples were run in quadruplicates.

### Protein-based surrogate neutralization ELISA

A protein-based surrogate neutralization ELISA was performed as described in (36). Essentially, 100 ng of purified RBD expressed in FreeStyle 293-F cells was immobilized overnight onto 96-well Immulon HBX plates (2 μg/ml), blocked, and incubated with four, 2-fold dilutions of patient samples, starting at 4 μl. Biotinylated ACE2 purified from FreeStyle 293-F cells was added (50 ng/well, incubated for 1hr), followed by streptavidin poly-HRP (Sigma, #S2438; 22 ng). 1-Step™ Ultra TMB-ELISA Substrate Solution (ThermoFisher, #34029) was added for 7.5 min at room temperature and the reaction was quenched with 50 μL stop solution containing 0.16 N sulfuric acid (ThermoFisher, #N600) and the optical density at 450 nm were read. The area under the curve of each dilution series for each patient plasma sample was calculated in R (version 4.0.1).

### Data and Statistical analysis

Flow cytometry data were analyzed using FlowJo v10. Multiplex cytokine bead data were analyzed using the LEGENDplex Data Analysis Software v8. All statistical and graphical analyses were performed using Graphpad Prism v6. Illustrations were created with Biorender.com. When data are shown in the absence of the control group, the values are calculated by subtracting background signal as indicated by “Δ” in panel labels. Background signal is defined by the frequency of cells expressing a particular cytokine, or concentration of an analyte, in wells cultured with BSA. The response was considered positive if the response to SARS-CoV-2 antigen was 10% higher than the response to BSA. For multiplex cytokine data, the limits of detection are indicated with dashed lines. Pair-wise comparisons were made by two-tailed Wilcoxon test, one-way ANOVA with Holm-Sidak’s multiple comparisons test or nonparametric Dunn’s multiple comparisons test as indicated in figure legends. Correlation analyses were performed by computing the Pearson or Spearman correlation coefficient. Statistical outliers were excluded from analyses by Grubb’s test, but all data points are displayed in figure panels.

## Data Availability

All data will be made available by the authors upon request, once paper has undergone full peer review and acceptance.

## Author contributions

JCL, WHK, PB, JL, KTA and BR performed experiments

FYY, JCL and MG processed patient samples

JL and JMR provided purified S protein,

AM, AC and MO recruited patients

SM provided SARS-CoV-2 virus

KTA, BR and AGC provided direct binding ELISA surrogate neutralization ELISA data JCL, WHK, PB, MO and THW designed the experiments and wrote the paper.

## Acknowledgments

We thank Birinder Ghumman for technical assistance, Payman Samavarchi-Tehrani, Derek Ceccarelli and Frank Sicheri, Mt. Sinai Hospital Toronto, for recombinant N protein purification and Jennifer Gommerman for helpful discussion. This research was funded by a FAST grant from the Thistledown foundation (to T.H.W.) and by a grant VR1-172711 from the Canadian Institutes of Health research to T.H.W., M.O. and A.C.G. M.O. receives funding from the Ontario HIV Treatment Network (OHTN), the Li Ka Shing Knowledge Institute, and the Juan and Stefania fund for COVID-19 and other virus infections. Funding for the development of the assays in the Gingras lab was provided through generous donations from the Royal Bank of Canada (RBC), QuestCap and the Krembil Foundation to the Sinai Health System Foundation; the equipment used is housed in the Network Biology Collaborative Centre at the Lunenfeld-Tanenbaum Research Institute, a facility supported by Canada Foundation for Innovation funding, by the Ontarian Government and by Genome Canada and Ontario Genomics (OGI-139). JCL and KTA were recipients of Ontario Graduate Scholarships.

## Supplemental Material

**Figure S1.**
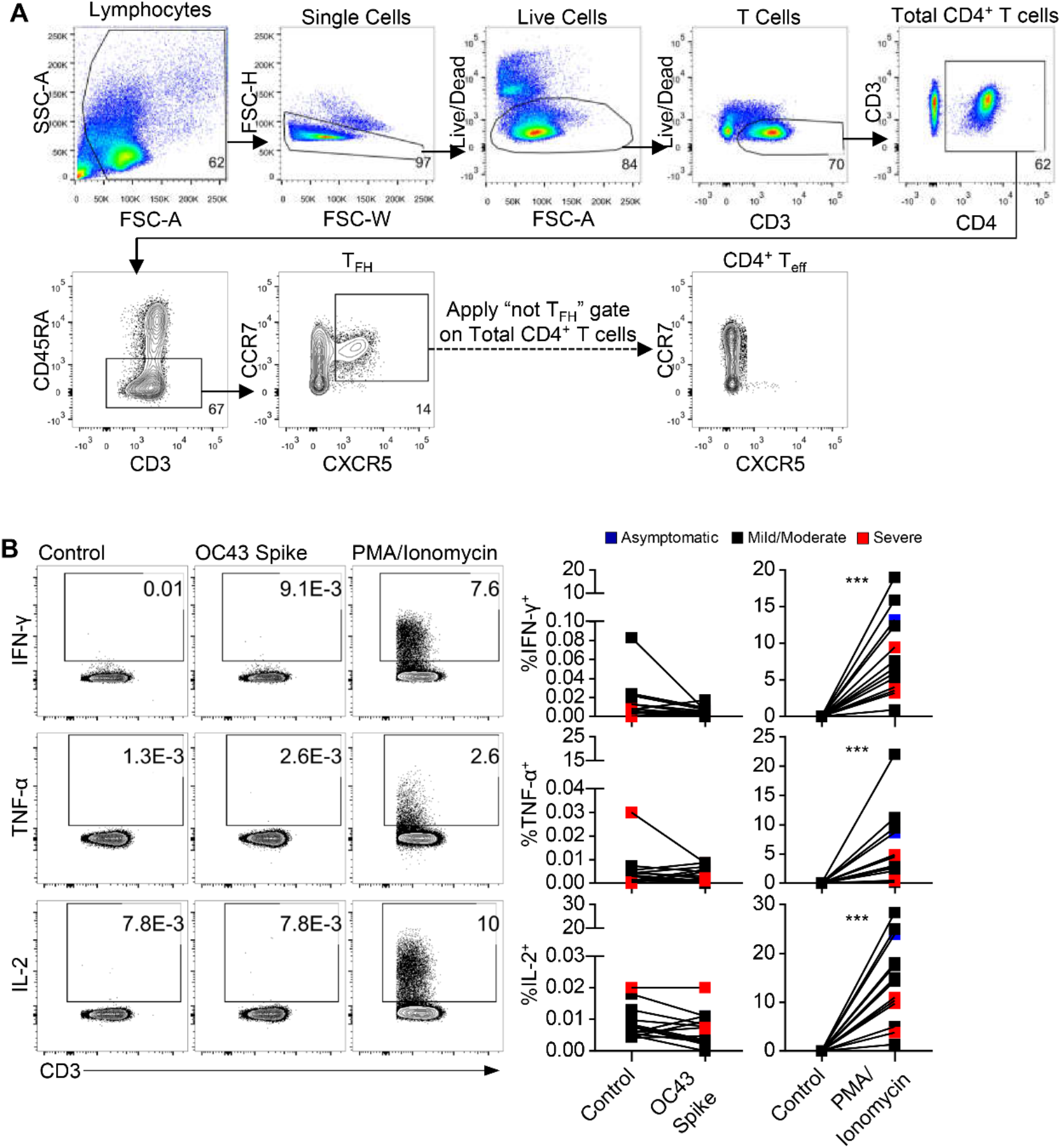
Gating strategy and controls. **(A)** Representative gating strategy for pTfh and non-pTfh CD4+ T cells. **(B)** Representative flow cytometry plots and graphs for CD4+ T cell IFN-γ, TNF-α, and IL-2 responses to OC43 S and PMA/Ionomycin (n=13), gated on non-pTfh CD4+ T cells. Pair-wise comparisons were made by two-tailed Wilcoxon test. ***p<0.001.

**Figure S2.**
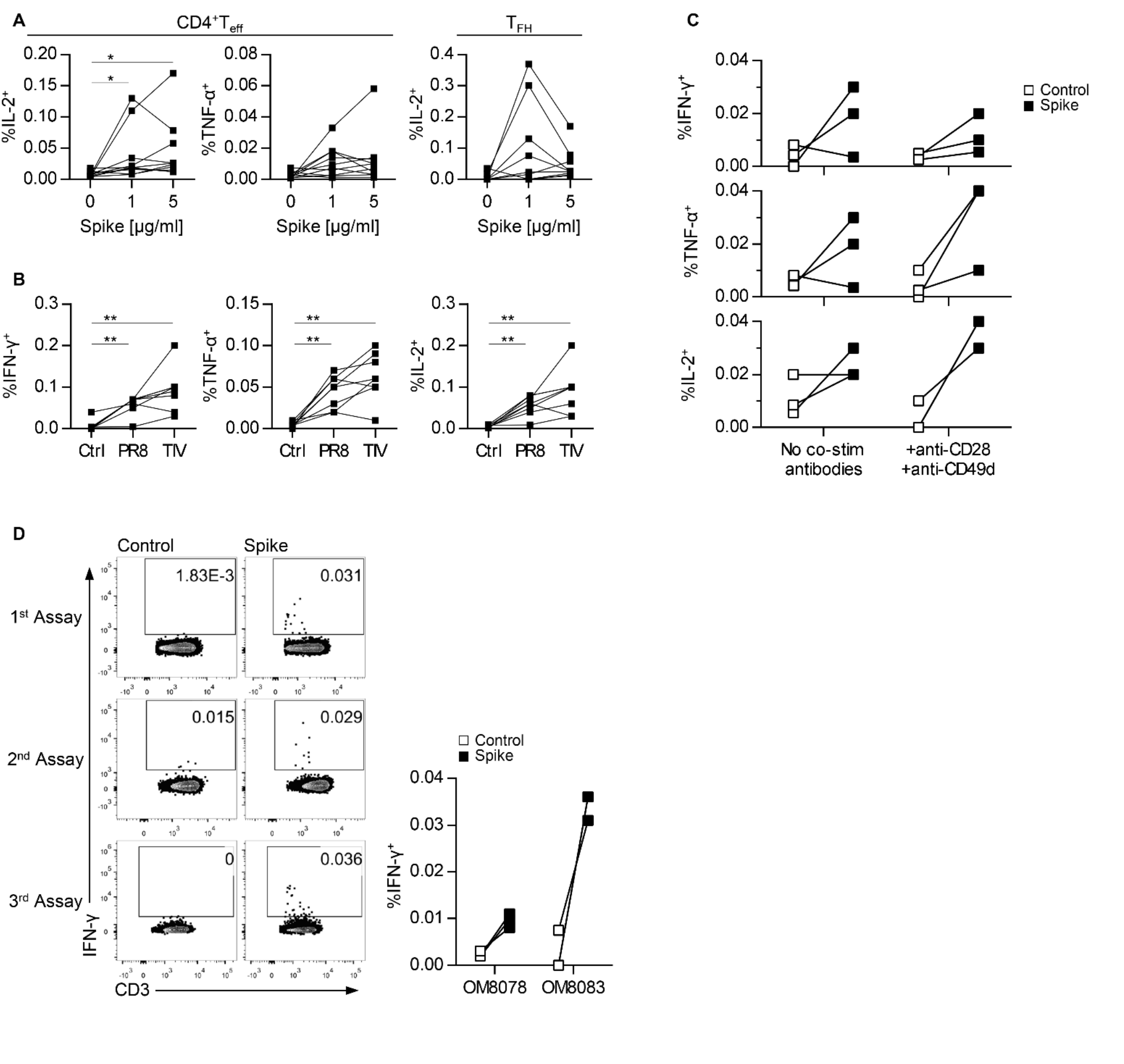
Determining optimal stimulation conditions. **(A)** CD4+ T cell IL-2 and TNF-α responses, and pTfh IL-2 responses, to two different concentrations of spike (n=10). **(B)** CD4+ T cell IFN-γ, TNF-α and IL-2 responses to PR8 compared to TIV (n=8). **(C)** CD4+ T cell IFN-γ, TNF-α and IL-2 responses to Spike with or without agonistic co-stimulatory antibodies anti-CD28 and anti-CD49d (n=3). **(D)** Representative flow cytometry plots and graphs showing pooled results from 3 independent experiments performed using PBMCs from the same group of donors to confirm assay reproducibility (n=2 per experiment). Nonparametric Dunn’s multiple comparisons test was performed for **(A)** and **(B)**. Two-way ANOVA was used to compare control and spike stimulated CD4+ T cells with or without co-stimulatory antibodies in **(C)**. *p<0.05, **p<0.01.

**Figure S3.**
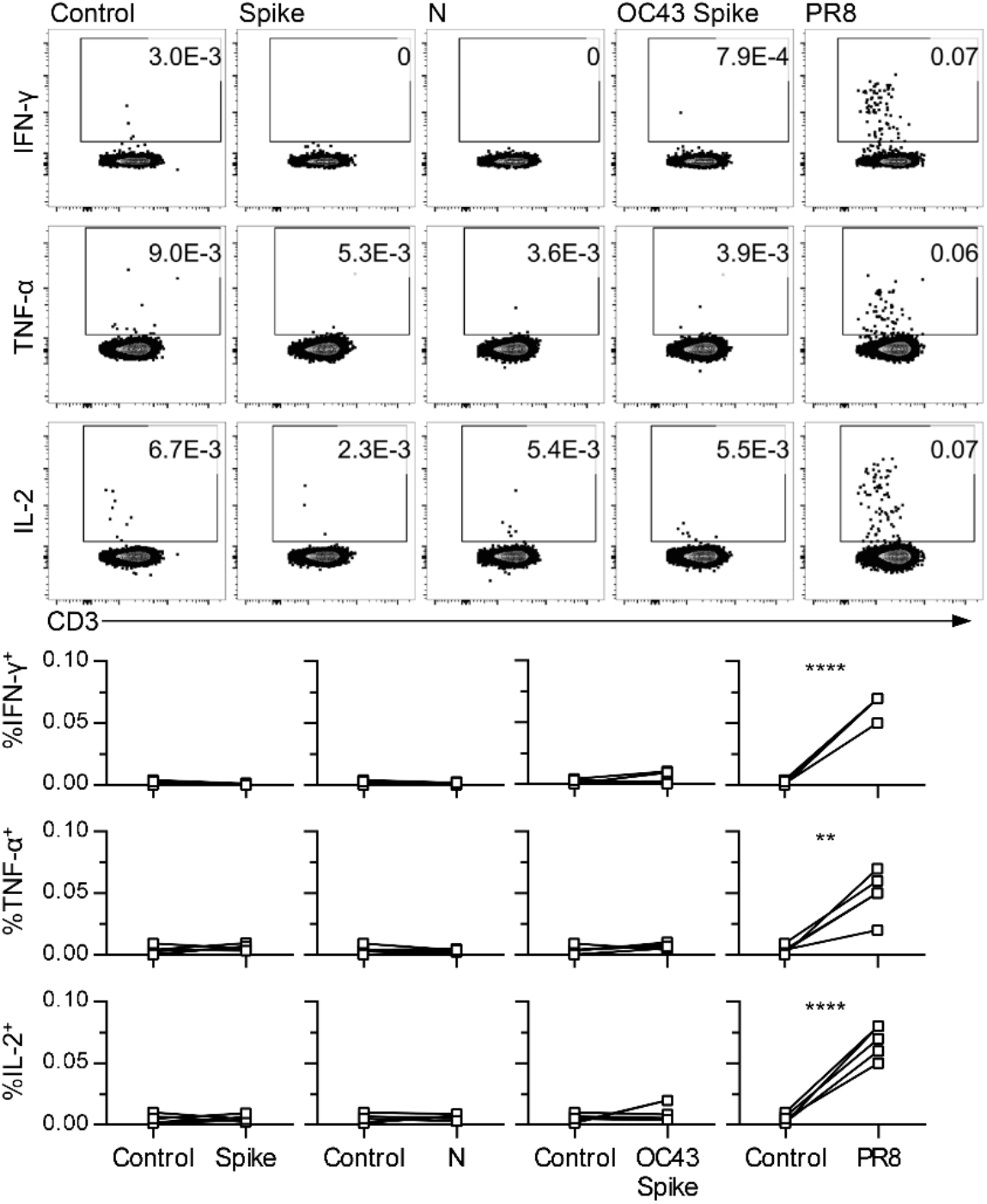
Healthy donor intracellular cytokine responses to viral antigen by flow cytometry. Representative flow cytometry plots and graphs for CD4+ T cell IFN-γ, TNF-α and IL-2 responses are shown in response to S, N, OC43 S or PR8 (n=5). Pair-wise comparisons were made by two-tailed Wilcoxon test. **p<0.01, ****p<0.0001.

**Figure S4.**
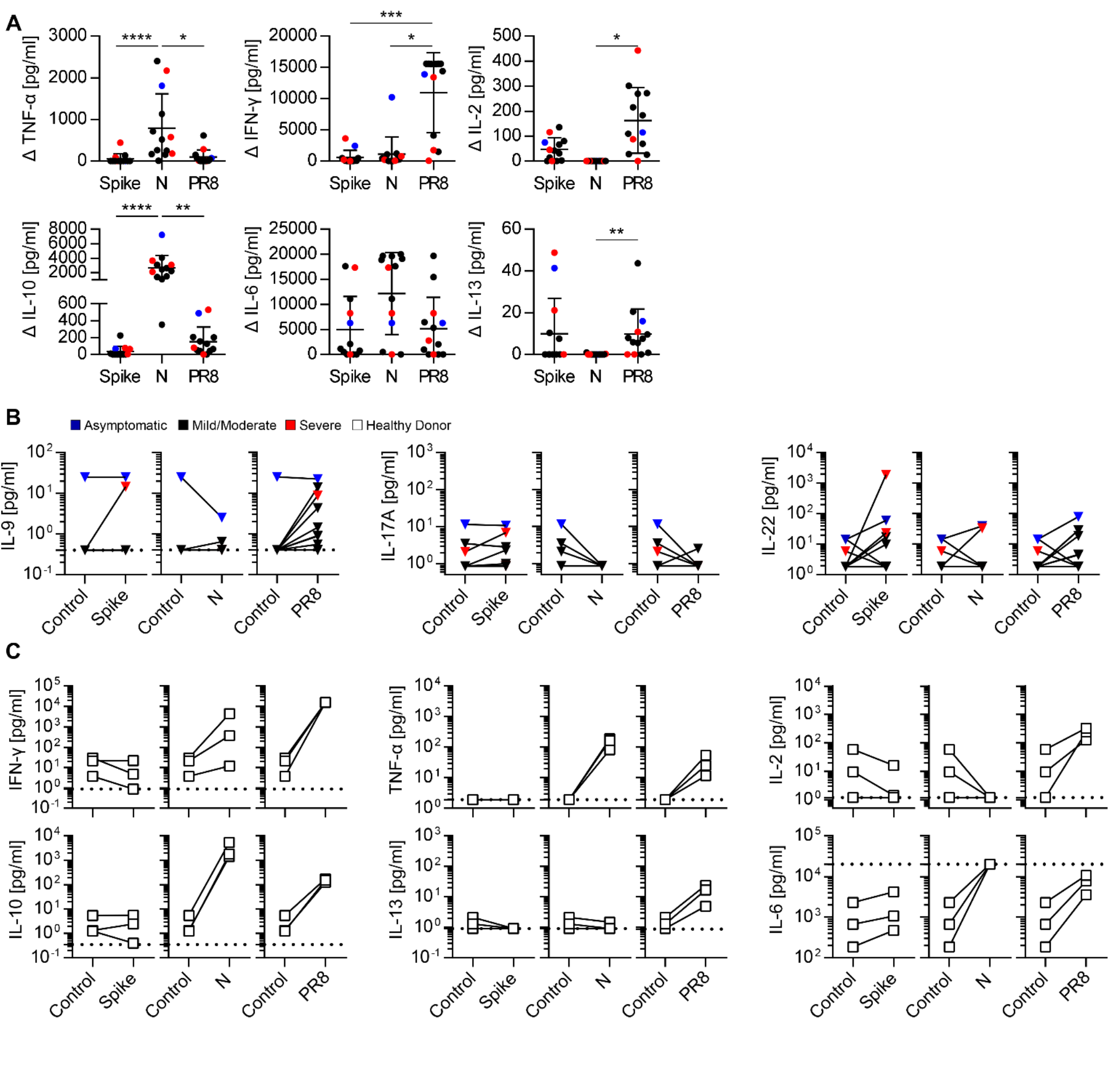
Cytokines secreted in response to PR8 and SARS-CoV-2 antigens. Graphs show levels of **(A)** TNF-α, IFN-γ, IL-2, IL-10, IL-6 and IL-13, **(B)** IL-9, IL-17A and IL-22 in SARS-CoV-2 convalescent PBMC cultures (n=13), and **(C)** levels of IFN-γ, TNF-α, IL-2, IL-10, IL-13 and IL-6 in healthy donor PBMC cultures (n=3) in response to S, N or PR8. Nonparametric Dunn’s multiple comparisons test was performed for **(A)**. Pair-wise comparisons were made by two-tailed Wilcoxon test in **(B)** and **(C)**. OM8099 exhibited high background TNF-α and was determined to be an outlier by the Grubb’s test. Although this data point is shown in all panels, it was excluded from statistical analysis of TNF-α responses. Graphs show mean±SD. *p<0.05, **p<0.01, ***p<0.001, ****p<0.0001.

**Figure S5.**
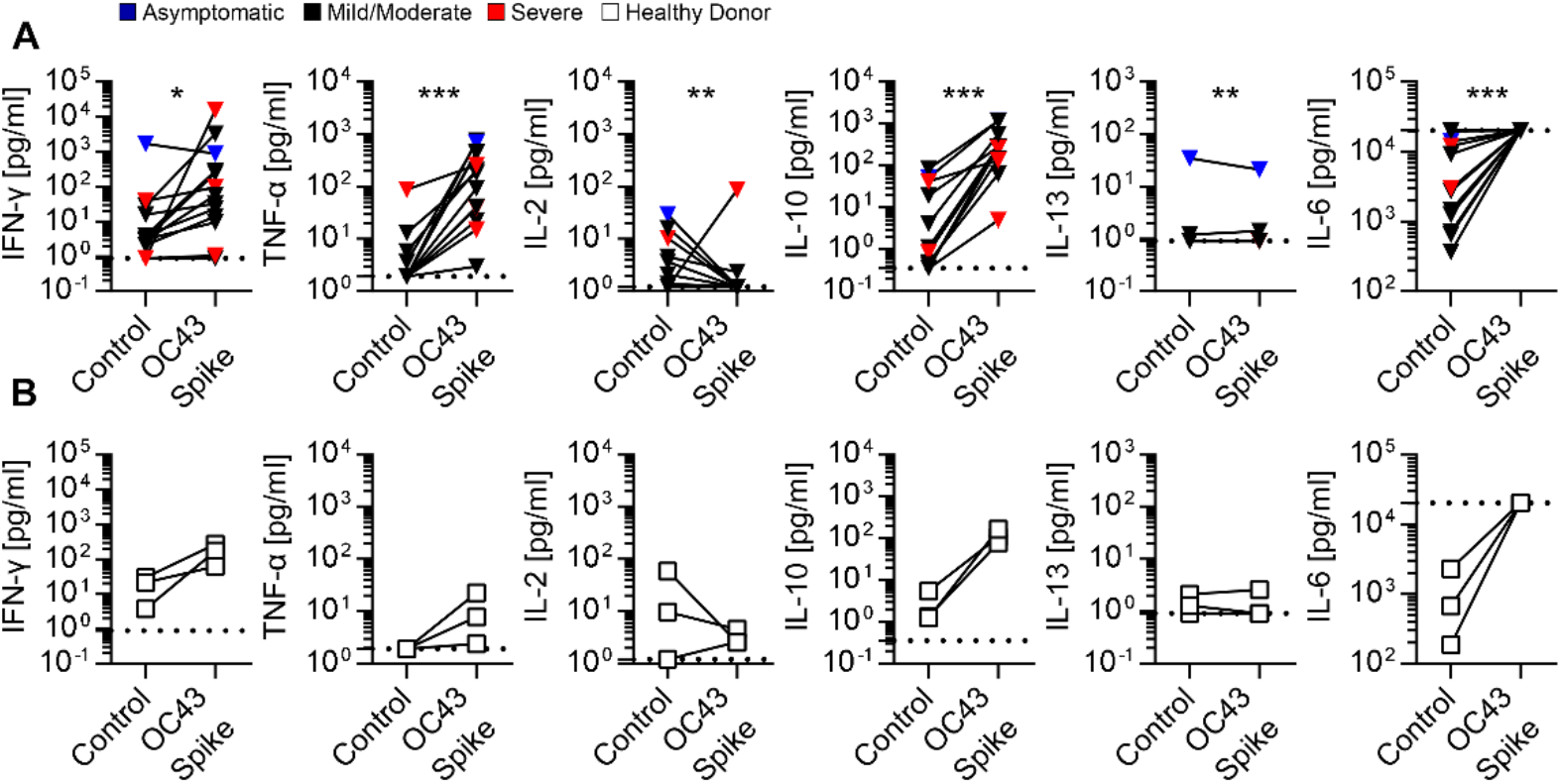
Cytokines secreted in response to OC43 S. Graphs show levels of IFN-γ, TNF-α, IL-2, IL-10, IL-13 and IL-6 in **(A)** SARS-CoV-2 convalescent PBMC cultures (n=13) and **(B)** healthy donor PBMC cultures (n=3) in response to OC43 S. Pair-wise comparisons were made by two-tailed Wilcoxon test. *p<0.05, **p<0.01, ***p<0.001.

**Figure S6.**
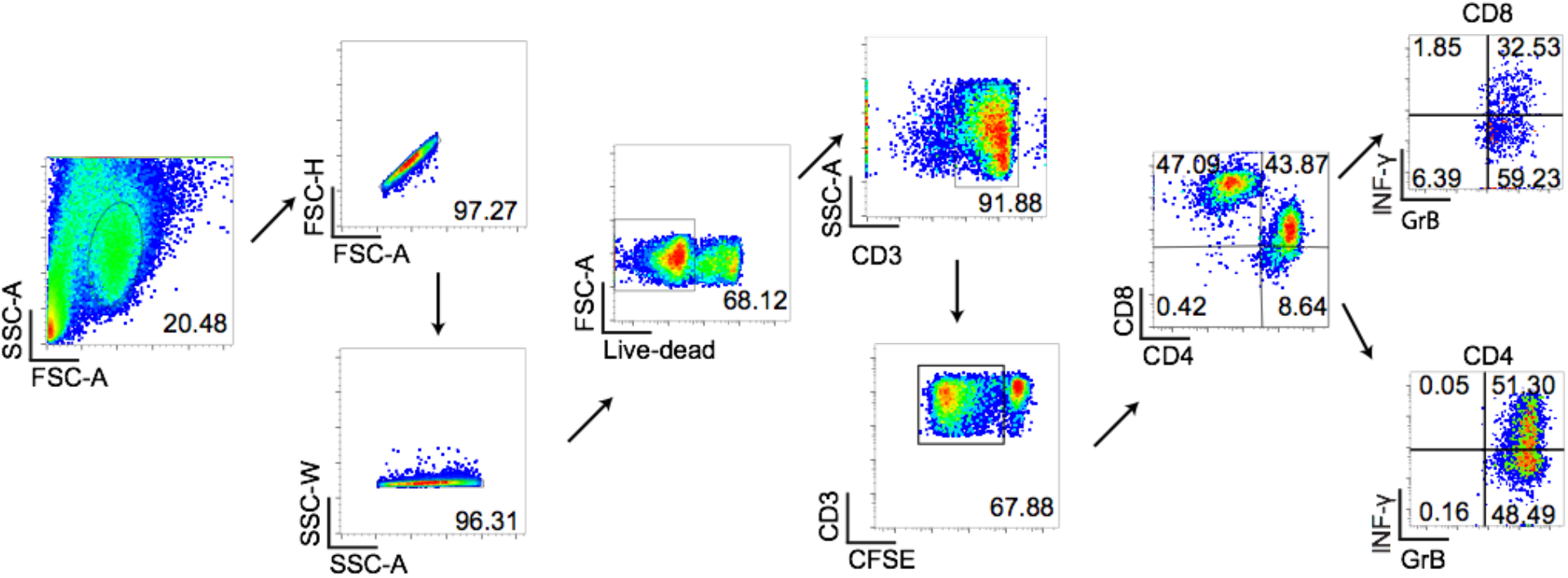
Flow cytometry gating strategy for T cells proliferative assay. Representative gating strategy for CFSE^low^ CD3+ T cells, CD4+/CD8+ T cells and IFN-γ/Granzyme B producing CD4+ /CD8+ T cells.

**Table S1**. *Summary of ICC, Multiplex Elisa and pTfh responses to S and N proteins and influenza A PR8 (excel file, uploaded separately)*.

**Table S2**. *Summary of T cell proliferation responses stimulated by four different COVID-19 master peptides pools (E, M, N, and S) for each patient (excel file, uploaded separately)*.

**Table S3.** - Antibody list.

| Antibody | Clone | Source | Catalog Number |
| --- | --- | --- | --- |
| Fixable Viability Dye eFluor™ 506 | N/A | eBiosciences | 65-0866-14 |
| anti-CD3 (AlexaFluor 700) | UCHT1 | BioLegend | 300424 |
| anti-CD4 (BUV395) | RPA-T4 | BD Biosciences | 564724 |
| anti-CD27 (BUV737) | L128 | BD Biosciences | 612829 |
| anti-CD45RA (BV786) | HI100 | BD Biosciences | 563870 |
| anti-CCR7 (BV605) | G043H7 | BioLegend | 353224 |
| anti-CXCR5 (BV421) | J252D4 | BioLegend | 356920 |
| anti-4-1BB (BV711) | 4B4-1 | BD Biosciences | 740798 |
| anti-HLA-DR (FITC) | L243 | BioLegend | 307604 |
| anti-IFN-γ (PE-Cy7) | 4S.B3 | BioLegend | 502528 |
| anti-TNF-α (PerCP/Cyanine5.5) | Mab11 | BioLegend | 502926 |
| anti-IL-2 (PE-eFluor 610) | MQ1-17H12 | eBiosciences | 61-7029-42 |
| anti-IL-17A (APC-eFluor 780) | eBio64DEC17 | eBiosciences | 47-7179-42 |
| anti-CD28 (Purified) | 9.3 | (In house) | N/A |
| anti-CD49d (Purified) | L25 | BD Biosciences | 340976 |
| anti-CD3 (APC-Cy7) | SK7 | BD Biosciences | 557382 |
| anti-CD4 (BV711) | SK3 | BD Biosciences | 563028 |
| anti-CD8 (PE) | HIT8a | BD Biosciences | 555635 |
| anti-IFNγ (APC) | 4S.B3 | BD Biosciences | 551385 |
| anti-granzyme B (BV421) | GB11 | BD Biosciences | 563389 |
| LIVE/DEAD™ fixable blue dead cell stain | N/A | Thermo Fisher Scientific | L23105 |
| Human TruStain FcX™ | N/A | BioLegend | 422301 |

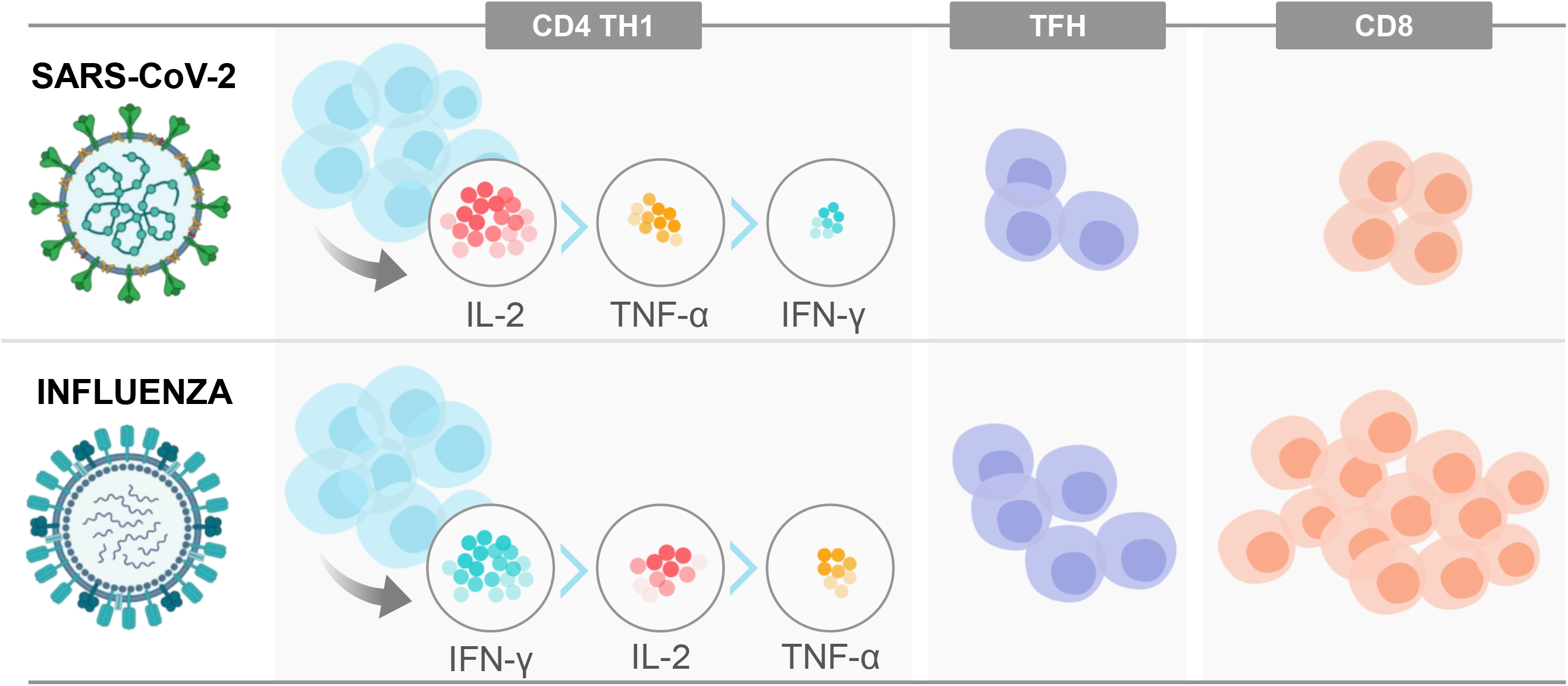

